# Recurrent dissemination of SARS-CoV-2 through the Uruguayan-Brazilian border

**DOI:** 10.1101/2021.01.06.20249026

**Authors:** Daiana Mir, Natalia Rego, Paola Cristina Resende, Fernando López-Tort, Tamara Fernandez-Calero, Verónica Noya, Mariana Brandes, Tania Possi, Mailen Arleo, Natalia Reyes, Matías Victoria, Andrés Lizasoain, Matías Castells, Leticia Maya, Matías Salvo, Tatiana Schäffer Gregianini, Marilda Tereza Mar da Rosa, Letícia Garay Martins, Cecilia Alonso, Yasser Vega, Cecilia Salazar, Ignacio Ferrés, Pablo Smircich, Jose Sotelo, Rafael Sebastián Fort, Cecilia Mathó, Ighor Arantes, Luciana Appolinario, Ana Carolina Mendonça, María José Benitez-Galeano, Martín Graña, Camila Simoes, Fernando Motta, Marilda Mendonça Siqueira, Gonzalo Bello, Rodney Colina, Lucía Spangenberg

## Abstract

**Background:** Uruguay is one of the few countries in the Americas that successfully contained the COVID-19 epidemic during the first half of 2020. Nevertheless, the intensive human mobility across the dry border with Brazil is a major challenge for public health authorities. We aimed to investigate the origin of SARS-CoV-2 strains detected in Uruguayan localities bordering Brazil as well as to measure the viral flux across this ∼1,100 km uninterrupted dry frontier.

**Methods:** Using complete SARS-CoV-2 genomes from the Uruguayan-Brazilian bordering region and phylogeographic analyses, we inferred the virus dissemination frequency between Brazil and Uruguay and characterized local outbreak dynamics during the first months (May-July) of the pandemic.

**Findings:** Phylogenetic analyses revealed multiple introductions of SARS-CoV-2 Brazilian lineages B.1.1.28 and B.1.1.33 into Uruguayan localities at the bordering region. The most probable sources of viral strains introduced to Uruguay were the Southeast Brazilian region and the state of Rio Grande do Sul. Some of the viral strains introduced in Uruguayan border localities between early May and mid-July were able to locally spread and originated the first outbreaks detected outside the metropolitan region. The viral lineages responsible for Uruguayan suburban outbreaks were defined by a set of between four and 11 mutations (synonymous and non-synonymous) respect to the ancestral B.1.1.28 and B.1.1.33 viruses that arose in Brazil, supporting the notion of a rapid genetic differentiation between SARS-CoV-2 subpopulations spreading in South America.

**Interpretation:** Although Uruguayan borders have remained essentially closed to non-Uruguayan citizens, the inevitable flow of people across the dry border with Brazil allowed the repeated entry of the virus into Uruguay and the subsequent emergence of local outbreaks in Uruguayan border localities. Implementation of coordinated bi-national surveillance systems are crucial to achieve an efficient control of the SARS-CoV-2 spread across this kind of highly permeable borderland regions around the world.

**Research in context:** *Evidence before this study:* Since the severe acute respiratory syndrome coronavirus 2 (SARS-CoV-2), causative agent of coronavirus disease 19 (COVID-19), was first detected in South America on February 26, 2020, it has rapidly spread through the region, causing nearly 350,000 deaths by December, 2020. In contrast to most American countries, Uruguay avoided an early exponential growth of SARS-CoV-2 cases and during the first six months of the pandemic it registered the lowest incidence of SARS-CoV-2 cases and deaths among South American countries. The intensive cross-border human mobility through the ∼1,100 km uninterrupted dry frontier between Uruguay and Brazil, might poses a major challenge for long-term control of the epidemic in Uruguay. Previous genomic studies conducted in Uruguay have analyzed sequences mostly sampled at the capital city, Montevideo, and detected prevalent SARS-CoV-2 lineages different from those described in Brazil, thus finding no evidence of frequent viral exchanges between these countries.

*Added value of this study:* Here we present the first genomic study of SARS-CoV-2 strains detected in different Uruguayan and Brazilian localities along the bordering region. The samples analyzed include 30% (*n* = 59) of all laboratory confirmed SARS-CoV-2 cases from Uruguayan departments at the Brazilian border between March and July, 2020, as well as 68 SARS-CoV-2 sequences from individuals diagnosed in the southernmost Brazilian state of Rio Grande do Sul between March and August, 2020. We demonstrate that SARS-CoV-2 viral lineages that widely spread in the Southeastern Brazilian region (B.1.1.28 and B.1.1.33) were also responsible for most viral infections in Rio Grande do Sul and neighboring Uruguayan localities. We further uncover that major outbreaks detected in Uruguayan localities bordering Brazil in May and June, 2020, were originated from two independent introduction events of the Brazilian SARS-CoV-2 lineage B.1.1.33, unlike previous outbreaks in the Uruguayan metropolitan region that were seeded by European SARS-CoV-2 lineages.

*Implications of all the available evidence:* Our findings confirm that although Uruguayan borders have remained essentially closed to non-Uruguayan citizens, dissemination of SARS-CoV-2 across the Uruguayan-Brazilian frontier was not fully suppressed and had the potential to ignite local transmission chains in Uruguay. These findings also highlight the relevance of implementing bi-national public health cooperation workforces combining epidemiologic and genomic data to monitor the viral spread throughout this kind of highly permeable dry frontiers around the world.

## INTRODUCTION

The severe acute respiratory syndrome coronavirus 2 (SARS-CoV-2), causative agent of coronavirus disease 19 (COVID-19), was first reported in South America on February 26, 2020, and rapidly spread through the region. South America is actually the second worst affected region in the world, with more than 11 million SARS-CoV-2 cases and nearly 350,000 deaths confirmed as of December, 2020^1^. While the virus exponentially spread during the first half of 2020 in most South American countries, the rapid implementation of non-pharmaceutical interventions avoided an exponential growth of SARS-CoV-2 cases in Uruguay^2,3^. Six months after the first four cases were reported on March 13, 2020, the country registered the lowest total and per capita numbers of SARS-CoV-2 cases (1,808 cases, 520 cases/million inhabitants) and deaths (45 deaths, 13 deaths/million inhabitants) in the region^4^.

Despite Uruguay’s success to control the early expansion of SARS-CoV-2, the intensive cross-border human mobility between Uruguay and neighboring countries heavily affected by the pandemic, is a major concern for public health authorities, aimed to achieve long-term epidemic control. With an area of approximately 176,000 km^2^ and 3.5 million inhabitants, Uruguay borders with Argentina to its west and southwest and with Brazil to its north and east. Of particular concern is the border with Brazil, a porous ∼1,100-kilometers strip of land that separates the southernmost Brazilian state of Rio Grande do Sul (RS) and the Uruguayan departments of Artigas (AR), Rivera (RI), Cerro Largo (CL), Treinta y Tres (TT) and Rocha (RO). The Brazilian-Uruguayan border hosts about 170,000 people that live in twin cities located both sides of a dry border and that maintain an intense economic and social interdependence^5^. With a total area of about 282,000 km^2^ and 11.3 million inhabitants, RS is the fifth-most-populous Brazilian state and as of July 31, 2020, registered 66,692 SARS-CoV-2 cases and 1,876 deaths^6^. After an initial phase of relatively slow growth, the COVID-19 epidemic displayed a sharp increase in RS since early May that coincides with the detection of several outbreaks along Uruguayan border departments. The largest SARS-CoV-2 Uruguayan outbreaks (∼50-100 confirmed cases) outside the metropolitan region during the first months after the first detected cases in the country, were at RI and TT departments in May and June, respectively, while smaller outbreaks (∼10-20 confirmed cases) were detected in AR and CL by July^7^. As of July 31, 2020, a total of 201 SARS-CoV-2 cases were reported in Uruguayan municipalities located along the border with Brazil, which represents 16% of the laboratory-confirmed cases in the country^7^.

Previous phylogenetic analyses revealed the circulation of different predominant SARS-CoV-2 lineages in Brazil (B.1.1.28 and B.1.1.33)^8,9^ and Uruguay (A.5 and B.1)^10,11^ thus supporting independent viral seeding events and little viral exchanges between these neighboring countries during the very early phase of the epidemic. However, most Uruguayan SARS-CoV-2 samples previously analyzed were from Montevideo, the capital city of the country and there is only scarce information concerning the virus strains circulating in Uruguayan localities bordering Brazil

We generated 122 SARS-CoV-2 whole genome sequences recovered from cases isolated at Uruguayan border departments (*n* = 54) as well as in the southernmost Brazilian state of RS (*n* = 68) to gain insight into the origin and dynamics of SARS-CoV-2 spread at the Brazilian-Uruguayan border. Our study provides important findings that demonstrate the relevance of building bi-national genomic surveillance workforces in countries with porous and dynamic bordering regions.

## METHODS

### SARS-CoV-2 samples and ethical aspects

A total of 122 SARS-CoV-2 whole-genomes were recovered from nasopharyngeal-throat combined swabs samples collected from deceased cases (n = 5), clinically ill or asymptomatic individuals that reside in five different Uruguayan departments (*n* = 54) at the Brazilian border and 41 different municipalities of the RS Brazilian state (*n* = 68) (Figure 1 and Table S1). Uruguayan samples were collected between May 5 and July 26, 2020, and underwent testing at the Universidad de la República, CENUR Litoral Norte, Salto (Molecular Virology Lab), Sanatorio Americano Montevideo (SASA, Molecular Biology Lab), Universidad de la República CURE Este, Rocha (Molecular Ecology Lab), and Laboratory of DILAVE/MGAP-INIA-UdelaR (Tacuarembó).

**Figure 1.**
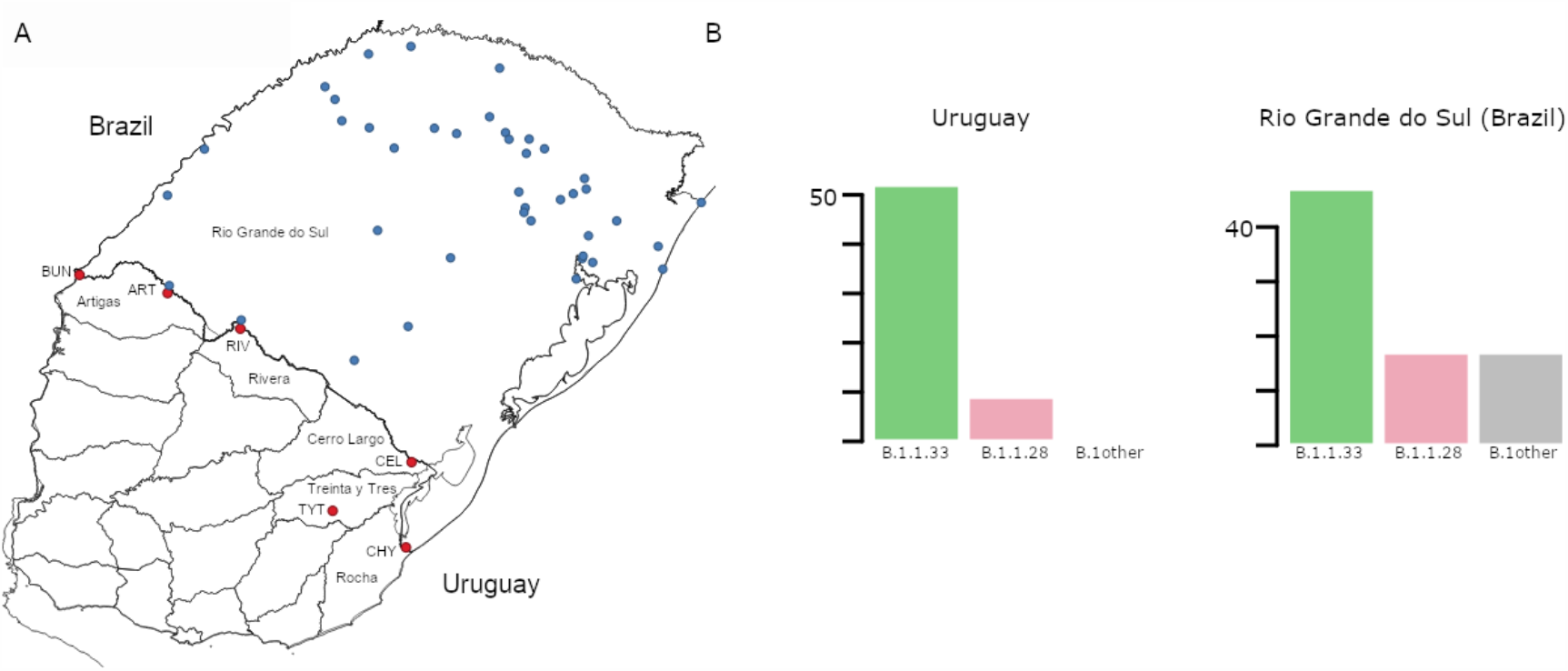
**A)** Map of Uruguay and Rio Grande do Sul (Brazil) showing the distribution of samples included in the study. Sampling locations in Uruguay and Rio Grande do Sul are marked in red and blue, respectively. Uruguayan departments bordering Brazil are explicitly named. Additionally, towns/cities within those departments are marked; BUN: Bella Unión, ART: Artigas, RIV: Rivera, CEL: Rio Branco, TYT: Treinta y Tres and CHY: Chuy. **B)** Prevalence of the different SARS-CoV-2 lineages detected in Uruguayan departments at the border region and in Rio Grande do Sul.

Sequencing of the Uruguayan samples was held at the Institut Pasteur de Montevideo (IPMON, Bioinformatics Unit) and Instituto de Investigaciones Biológicas Clemente Estable (IIBCE, Genomics Department). Brazilian samples were recovered from sentinel locations in RS state from March 9 to August 16, 2020, and sent to the central laboratory from RS state (LACEN-RS) for SARS-CoV-2 RT-PCR detection. Positive Brazilian samples were subsequently sent to the Laboratory of Respiratory Viruses and Measles, IOC, FIOCRUZ, WHO Regional Reference Laboratory for Coronavirus in the Americas. This study was approved by Ethics Committees in Uruguay (SASA Ethics Committee; supplementary materials) and Brazil (FIOCRUZ-IOC Ethics Committee: 68118417.6.0000.5248, and Brazilian Ministry of Health SISGEN: A1767C3).

### SARS-CoV-2 identification of positive samples, amplification and sequencing

Molecular detection of the virus was performed with different kits at each center (Biomanguinhos SARS-CoV-2 kit, OneStep RT-qPCR kit COVID-19 RT-PCR Real TM Fast, Coronavirus COVID-19 genesig® Real-Time PCR assay), according to manufacturer’s instructions (supplementary materials). SARS-CoV-2 genomes were recovered using both long and short PCR amplicon protocols^12,13,14^; and three sequencing technologies: Oxford Nanopore Technologies (ONT), Illumina and Ion Torrent (Table S1). See supplementary methods for details.

### SARS-CoV-2 whole-genome consensus sequences

Whole-genome consensus sequences obtained from ONT were generated using an adaptation of the nCoV-2019 novel coronavirus Artic bioinformatics protocol (https://artic.network/ncov-2019/ncov2019-bioinformatics-sop.html) as in^15^ with adjustments that are available at https://github.com/iferres/ncov2019-artic-nf. For Illumina and Ion Torrent, bcftools was used for SNP calling (mpileup), SNP filtering and consensus sequences reconstruction (consensus function) as described in detail in supplementary material. Positions of interest were manually inspected to resolve undetermined bases. All genomes obtained in this study were uploaded at the EpiCoV database in the GISAID initiative under the accession numbers EPI_ISL_729794 to EPI_ISL_729861 (Brazilian genomes) and accession numbers of Uruguayan genomes are in Table S2.

### SARS-CoV-2 genotyping and Maximum Likelihood phylogenetic analyses

Uruguayan and Brazilian SARS-CoV-2 genome sequences were initially assigned to viral lineages according to^16^, using the Pangolin web application (https://pangolin.cog-uk.io) and later confirmed using maximum likelihood (ML) phylogenetic analyses. ML phylogenetic analyses were performed with the PhyML 3.0 program^17^, using an online web server^18^ (supplementary materials). Branch support was assessed by the approximate likelihood-ratio test based on a Shimodaira–Hasegawa-like procedure (SH-aLRT) with 1,000 replicates^19^.

### Phylogeographic analyses

The previously generated ML trees were employed for the ancestral character state reconstruction (ACR) of epidemic locations with PastML^20^, using the Marginal Posterior Probabilities Approximation (MPPA) method with an F81-like model. We next constructed a time-scaled Bayesian phylogenetic tree for the Brazilian and Uruguayan sequences belonging to the B.1.1.28 and B.1.1.33 lineages using the Bayesian Markov Chain Monte Carlo (MCMC) approach implemented in BEAST 1.10^21^ with BEAGLE library v3 to improve computational time (supplementary materials). Viral migrations were reconstructed using a reversible discrete phylogeographic model^22^ with a continuous-time Markov chain (CTMC) rate reference prior^23^.

### Within-host diversity

Only those synapomorphic sites with at least 100 reads were kept for further analysis. As samples were obtained by three different sequencing technologies with different error profiles, a Shannon Entropy value (H) was estimated per observation based on the four allele frequencies. We determined a linear model to estimate the contributions of sequencing technology, sample and synapomorphic site, to the observed A, C, G, T frequencies (summarized as H): H exp ¼ ∼ SEQ + SAMPLE + MUTATION. We used H exp ¼ to better fit a normal distribution (Figure S1C). Details in supplementary materials.

## RESULTS

### Prevalent SARS-CoV-2 lineages at the Uruguayan-Brazilian border

To understand the dynamics of SARS-CoV-2 spread at the Brazilian-Uruguayan border, we sequenced the viral genome from 54 individuals diagnosed between May 5 and July 27 in the five Uruguayan border departments (AR, CL, RI, RO and TT) and from 68 individuals diagnosed at 42 different municipalities from RS state collected between March 9 and August 16, 2020 (Figure 1A and Table S1). These sequences were combined with a few SARS-CoV-2 whole-genomes sequences from Uruguayan individuals diagnosed at RI (n = 5) and Montevideo (n=1), and from Brazilian individuals sampled in RS (n = 13) retrieved from the EpiCoV database in the GISAID initiative. The resulting final dataset of 59 SARS-CoV-2 sequences from Uruguayan departments bordering Brazil represents 30% of all laboratory-confirmed cases (*n* = 201) in that region between March and July, 2020. A low fraction of Uruguayan (∼4%) and no Brazilian individuals here sequenced reported international travel or contact with traveling people, indicating that most of them were locally infected. The SARS-CoV-2 genotyping of Uruguayan and Brazilian diagnosed at the border region revealed a quite homogenous pattern as all sequences belonged to the B.1 lineage characterized by the D614G mutation at the spike protein. Furthermore, all SARS-CoV-2 sequences from Uruguayan departments at the border region belonged to the dominant Brazilian lineages B.1.1.33 (85%) and B.1.1.28 (15%) (Figure 1B). Although different B.1 sub-clades were identified in RS, the Brazilian lineages B.1.1.33 (58%) and B.1.1.28 (21%) were also the most prevalent across all state regions (Figure 1B).

### Identification of major SARS-CoV-2 Uruguayan-Brazilian clades

Phylogenetic trees of SARS-CoV-2 lineages B.1.1.28 and B.1.1.33 were inferred to explore the Uruguayan and Brazilian-RS sequence clustering with sequences from other Brazilian states and worldwide. The ML phylogenetic analyses revealed that most SARS-CoV-2 sequences from Uruguay branched within four highly supported (aLRT = 1) Uruguayan clades (Figure 2). The largest B.1.1.33 Uruguayan clade TT-I_33_ (n = 28) comprises all sequences detected in TT between June 18 and July 2. The B.1.1.33 clades RI-I_33_ (n = 19) and RI-II_33_ (n = 3) comprises all sequences detected in RI between May 5 and June 5 and in late July, respectively (Figure 2A). The B.1.1.28 Uruguayan clade AR-I_28_ (n = 4) comprises all sequences detected in AR (Bella Unión city) in late July (Figure 2B). The B.1.1.33 sequence detected in March in Montevideo as well as the B.1.1.28 sequences detected in July in AR (Artigas city), CL, RO and RI, appeared as dyads or singletons.

**Figure 2.**
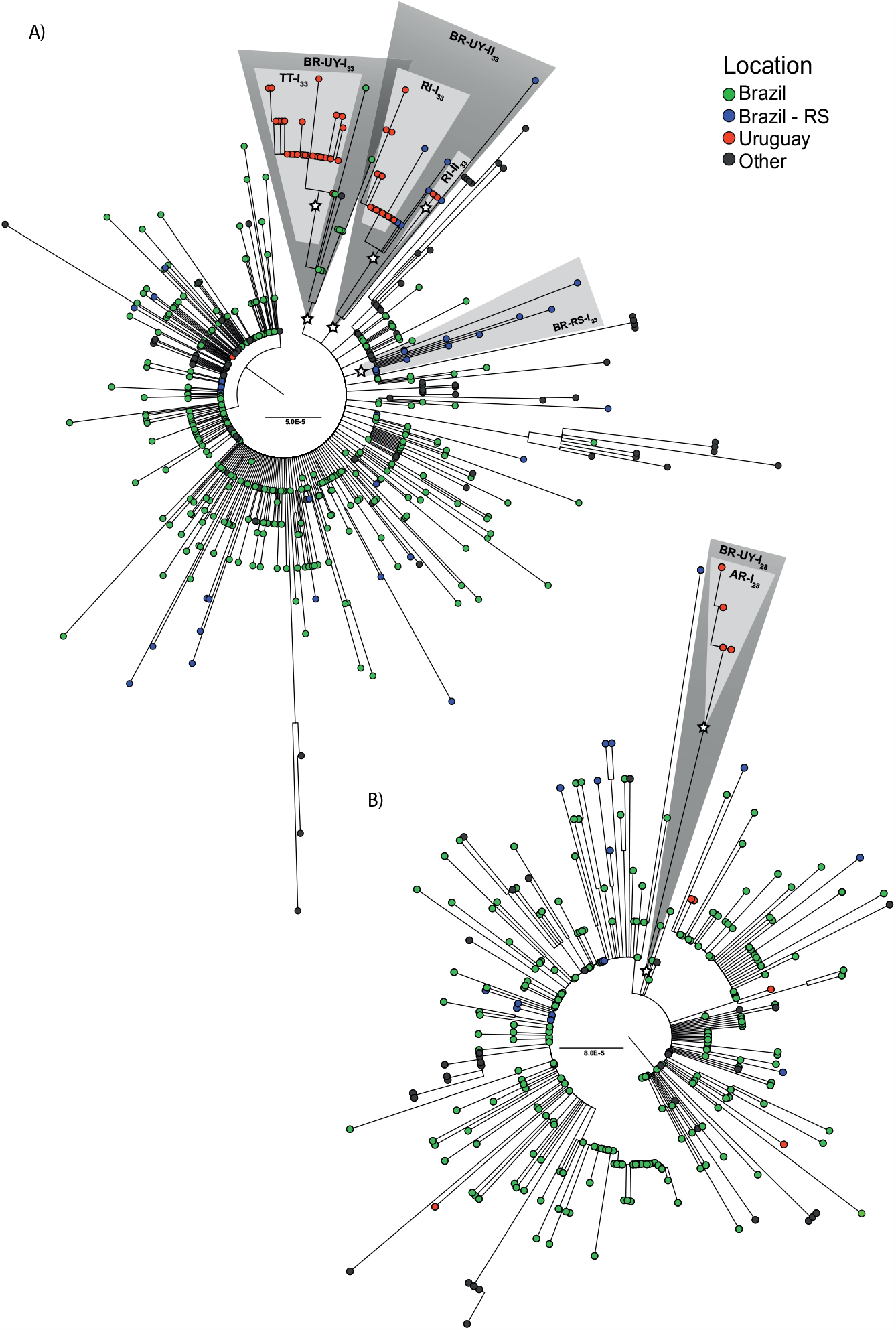
Identification of major SARS-CoV-2 Uruguayan-Brazilian clades. ML phylogenetic trees of **A)** 86 B.1.1.33 and **B)** 23 B.1.1.28 genomes obtained in this study along with 492 and 275 worldwide reference sequences of the respective genotypes available in GISAID database. Tip circles are colored according to the sampling location. Node supports (aLRT) values at key nodes are represented by (*). Shaded boxes highlight the position of clusters BR-UY-I_33,_ BR-UY-II_33,_ BR-RS-I_33,_ BR-UY-I_28,_ TT-I_33,_ RI-I_33,_ RI-II_33_ and AR-I_28._ The tree was rooted on midpoint and branch lengths are drawn to scale with the bars at the center indicating nucleotide substitutions per site. UY-AR: Artigas-Uruguay, UY-RI: Rivera-Uruguay, UY-TT: Treinta y Tres-Uruguay, BR-SE: Southeast Brazilian region, BR-RS: Rio Grande do Sul-Brazil.

The four major Uruguayan clades were nested among basal sequences from Brazil, forming three highly supported (aLRT = 1) Brazilian-Uruguayan clades (Figure 2). The largest clade (n = 39) designated as BR-UY-I_33_ comprises the Uruguayan clade TT-I_33_, a group of basal Brazilian sequences (n = 10) isolated in São Paulo and Rio de Janeiro states between March 24 and June 6, and one sequence from Ireland. The second Brazilian-Uruguayan clade designated as BR-UY-II_33_ comprises the Uruguayan clades RI-I_33_ (n = 19) and RI-II_33_ (n = 3) and a group of Brazilian sequences (n = 8) isolated in different municipalities from RS between May 6 and July 28 (Figure 2A). The third Brazilian-Uruguayan clade designated as BR-UY-I_28_ comprises the Uruguayan clade AR-I_28_ and one Brazilian sequence isolated in the São Paulo state on March 20 (Figure 2B). Our analysis also identified one highly supported (aLRT = 1) monophyletic group designated as BR-RS-I_33_ (n = 12) that only comprises sequences from RS (Figure 2A).

### Spatiotemporal dissemination of SARS-CoV-2 Uruguayan-Brazilian clades

To identify the number of independent introduction events of SARS-CoV-2 into Uruguay and their most probable source location we used a ML-based probabilistic method of ancestral character states reconstruction implemented in the PastML program. Sequences were grouped according to country (Argentina, Brazil, Chile and Uruguay) or region (North America, Europe and Oceania) of origin. The ML phylogeographic analyses estimate nine separate viral introductions into Uruguay from Brazil: five of the lineage B.1.1.28 and four of the lineage B.1.1.33 (Figure 3). Three introductions of lineage B.1.1.33 and one introduction of lineage B.1.1.28 led to onward transmission to more than one individual and gave origin to the Uruguayan clades RI-I_33_, RI-II_33_, TT-I_33_ and AR-I_28_.

**Figure 3.**
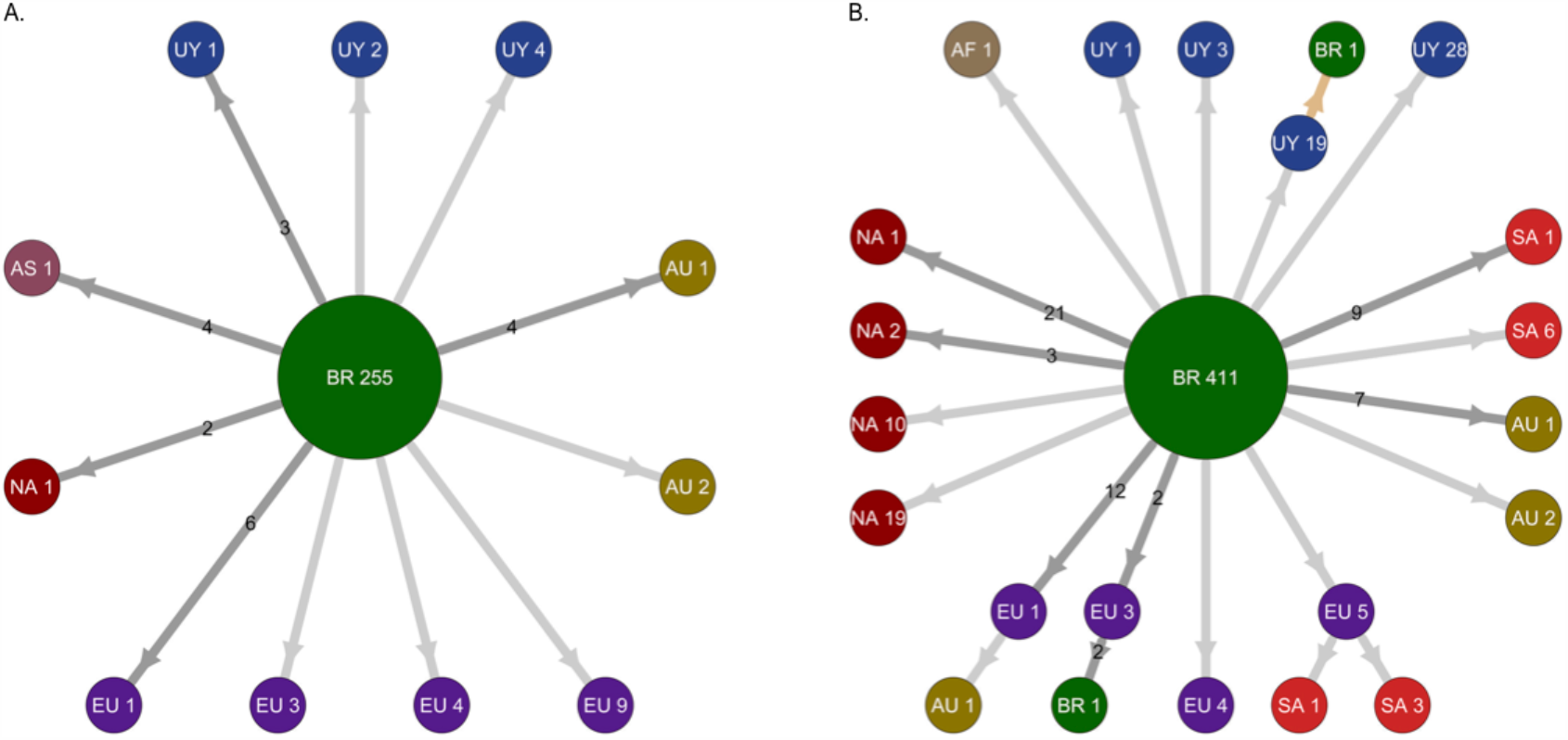
Migration events during worldwide dissemination of SARS-CoV-2 lineages B.1.1.28 and B.1.1.33. The picture depicts the migration events in SARS-CoV-2 lineages B.1.1.28 **A)** (*n* = 298 sequences) and B.1.1.33 **B)** (*n* = 578 sequences) inferred by ancestral character reconstruction obtained through a ML method implemented in PastML. Each node in the network is associated with a location and accompanied by the number of sequences connected by neighboring nodes with similar reconstructed location. Reiterated migration events are represented by links in which its number of occurrences are indicated. The light brown arrow points out an event inferred only through ML methods. AF: Africa, AS: Asia, AU: Australia, BR: Brazil, EU: Europe, NA: North America, SA: South America, UY: Uruguay.

To more accurately infer the geographic source and timing of virus introductions in Uruguay, we conducted Bayesian-based phylogeographic and molecular clock analyses of Brazilian and Uruguayan sequences. Viral migrations were inferred using a discrete diffusion model between 13 locations: four Brazilian regions (Southeast, Northeast, North and Central-West), three southern Brazilian states (RS, Parana and Santa Catarina), and six Uruguayan locations (AR, CL, MO, TT, RI and RO). Bayesian analyses recovered the same Uruguayan and Brazilian clades previously detected by the ML analyses, with two exceptions: the clade RI-II_33_ branched outside the clade BR-UY-II_33_ and no Brazilian sequences branched with high support with clade AR-II_28_. Bayesian analyses pointed that the Southeast region was the most probable source (posterior state probability [*PSP*] ≥ 0.96) of all B.1.1.28 and B.1.1.33 viruses introduced in Uruguay, with exception of clade RI-I_33_, a B.1.1.28 sequence from RI that most probably originated in Brazil-RS (*PSP* ≥ 0.85) and the B.1.1.33 strain from Montevideo that probably originated in the North Brazilian region (*PSP* = 1) (Figure 4 and Figures S2 and S3).

**Figure 4.**
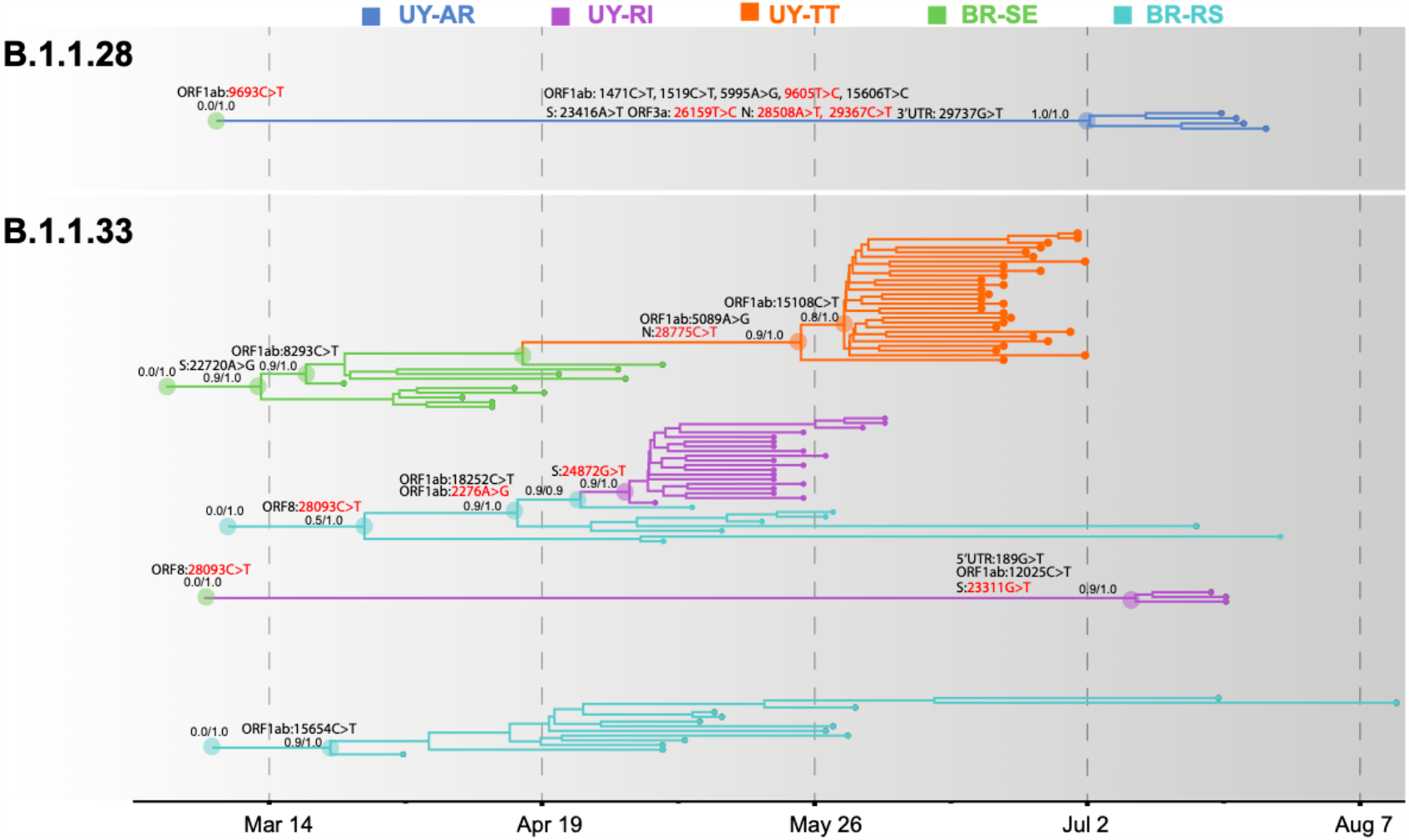
Spatiotemporal dissemination of SARS-CoV-2 Uruguayan-Brazilian clades. Uruguayan-Brazilian clades inferred on the time-scaled bayesian phylogeographic MCC tree (Figures S2 and S3) plotted independently. Branches are colored according to the most probable location state of their descendant nodes as indicated at the legend. Posterior probability/Posterior state probability support values are indicated at key nodes. Synonymous (black) and non-synonymous (red) substitutions fixed at ancestral nodes are shown.

The temporal origin of the lineages B.1.1.28 and B.1.1.33 was traced back to February 2020 and the T_MRCA_ of major Brazilian-Uruguayan clades were inferred at: March 12 (95% HPD: March 09-21) for clade BR-UY-I_33_, March 23 (95% HPD: March 14-31) for clade BR-RS-I_33_, March 29 (95% HPD: March 12 - April 13) for clade BR-UY-II_33_, May 2 (95% HPD: April 27 - May 04) for clade RI-I_33_, May 26 (95% HPD: May 12 - June 07) for clade TT-I_33_, July 10 (95% HPD: June 27 - July 18) for clade RI-II_33_, and July 2 (95% HPD: June 17 - July 16) for clade AR-I_28_. The detection lag (time interval between cluster T_MRCA_ and the first detected case) of major B.1.1.33 Uruguayan outbreaks was estimated at 3 days (95% HPD: 1-8 days) for clade RI-I_33_ and 23 days (95% HPD: 11-37 days) for clade TT-I_33_. The control lag (time interval between the first detected case and the last transmission event) was estimated at 25 days (95% HPD: 20-28 days) for clade RI-I_33_ and 10 days (95% HPD: 6-12 days) for clade TT-I_33_.

### SARS-CoV-2 lineage-defining SNPs and intra-patient viral diversity

SARS-CoV-2 Brazilian-Uruguayan clades here identified displayed several lineage-defining SNPs (single nucleotide polymorphisms/mutations) that were sequentially fixed during evolution and dissemination of each lineage (Figure 4 and Table S3). The Uruguayan/Brazilian lineages (BR-UY-I_33_, BR-UY-II_33_ and BR-UY-I_28_) and the Brazilian lineage BR-RS-I_33_ were all defined by one SNP. One SNP also characterized all sequences from clade TT-I_33_ plus some basal Brazilian strains from São Paulo, two SNPs defined clade TT-I_33_ and one additional SNP characterized all except one basal sequence of clade TT-I_33_. Two SNPs characterized all sequences from clade RI-I_33_ plus some basal Brazilian strains from RS, one SNP defined clade RI-I_33_ and three SNPs defined clade RI-II_33_. The clade AR-I_28_ was defined by 10 SNPs (Table S3).

To better understand the dynamics of lineage-defining SNPs, we examine within-host diversity patterns by analyzing the mapped reads at every SNPs position along the genome in SARS-CoV-2 samples from the two major B.1.1.33 Brazilian-Uruguayan clades (Table S4, S5). Our analysis of SNPs positions reveals that mutant alleles displayed either low (<10%) or high (> 80%) frequency within each clade and we found no evidence of sequences with intermediate frequencies (10-65%) (Figure 5A). The most notable example was the mutation ORF1ab:15108C>T fixed during the dissemination of clade TT-I_33_ that appeared at a very low frequency (<5%) in the basal Uruguayan sample M70 (Figure 5B). Indeed, the estimated frequency of the mutant allele in basal sequences from a given clade was not higher than the corresponding frequency detected in sequences from a different clade (Figure 5C).

**Figure 5.**
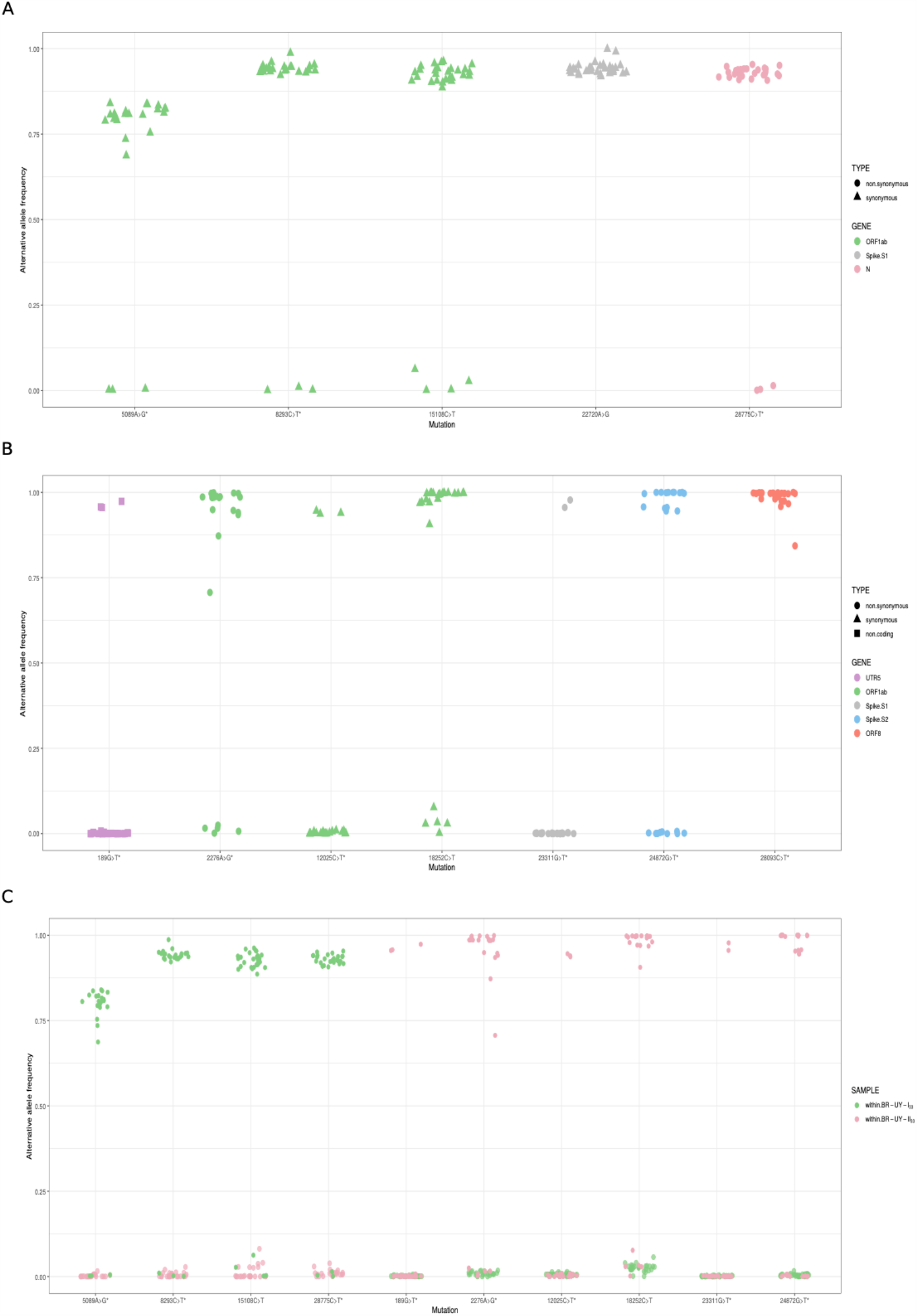
Within host-diversity in synapomorphic sites for the SARS-CoV-2 clades BR-UY-I_33_ and BR-UY-II_33_. **A**) Alternative allele frequencies for the clade BR-UY-I_33_, as observed from trimmed bam files, are shown. Positions and annotation follow Wuhan’s reference sequence MN908947; corresponding gene annotation and mutation types are indicated by color and shape, respectively. The asterisk indicates the synapomorphic site where the MUTATION explanatory variable shows a significant coefficient in the implemented linear model (p-value <= 0.05). In other words, the allele frequencies for this site are robust to sequencing technology. **B**) Same as A, but for the clade BR-UY-II_33_. For both clades, the absence of alleles with intermediate frequencies (0.1 < f < .65) is notorious. **C**) Alternative allele frequencies for all synapomorphies within both BR-UY-I_33_ and BR-UY-II_33_ clades. For each mutation, there is overlap in the alternative allele frequencies of those samples that do not show the synapomorphy, regardless of the clade they belong to.

## DISCUSSION

Public health measures implemented by authorities were able to contain the early local expansion of SARS-CoV-2 in Uruguay, but the intensive human mobility across the contiguous dry borderland with Brazil can compromise long-term control of the epidemic. Indeed, our analyses showed that the COVID-19 epidemic in the Uruguayan-Brazilian border is mostly dominated by the Brazilian lineages B.1.1.33 and B.1.1.28 and that independent introductions of these lineages seeded the early outbreaks of SARS-CoV-2 detected in Uruguayan departments at the bordering region. Our phylogeographic analyses support that both SARS-CoV-2 lineages most probably arose in the Southeast Brazilian region (*PSP* = 1) during February 2020, consistent with previous findings^8,15^ and were later disseminated at multiple times into the Southern region, making up 79% of all viral strains from the RS state. We distinguished two B.1.1.33 local sub-clades that together comprise 45% of all B.1.1.33 strains from RS: the clade BR-RS-I_33_ that arose by the end of March and seems to have remained restricted to this Brazilian state, and the clade BR-UY-II_33_ that arose between late March and early April and was later disseminated to Uruguay. These findings indicate that the COVID-19 epidemic in the Southern Brazilian state of RS was mostly seeded by epidemics from the Southeastern region.

The molecular epidemiologic profile of the COVID-19 epidemic at Uruguayan border localities (dominated by lineages B.1.1.33 and B.1.1.28) coincides with the observed pattern in RS state and differs from the profile of early outbreaks in the capital city of Montevideo (dominated by lineages A.5 and B.1)^10,11^. Our phylogeographic analyses support that the Southeast Brazilian region and RS were the most probable sources of seven and two viral introductions, respectively, into Uruguay. This finding, however, should be interpreted with caution because potential sampling bias across Brazilian regions might influence phylogeographic reconstructions and the actual number of viral introductions from RS into Uruguay might be higher than here estimated^24^.

Our study supports that some of the viral strains introduced in the Uruguayan cities of Rivera, Treinta y Tres and Bella Unión between early May and mid-July were able to spread locally. The most successfully disseminated local lineages were RI-I_33_ and TT-I_33_ that drove the major early outbreaks detected in Rivera and Treinta y Tres in May and June, respectively. Our analysis supports that SARS-CoV-2 Uruguayan clades displayed a variable period of local cryptic transmission, lasted between one and three weeks. Furthermore, the major Uruguayan clades took between 10 days and nearly a month to be controlled. Long periods of cryptic circulation and/or long times to control, represent a potential risk of viral dissemination from Uruguayan border localities into the metropolitan region.

Our phylogenetic analyses identified several sub-clusters inside the B.1.1.28 and B.1.1.33 lineages defined by a set of synonymous and non-synonymous mutations with respect to the ancestral viruses that arose in Brazil in February, 2020. The number of defining mutations ranges from one to 11 SNPs and correlates with the lineage T_MRCA_ as lineages with the youngest T_MRCA_ displayed more SNPs than those with older ones. This is consistent with a molecular clock model^25^ and with the notion that most SNPs are probably evolutionary neutral^26^. Thus, while our study emphasizes the rapid genetic differentiation of regional SARS-CoV-2 lineages in South America and the relevance of molecular epidemiologic studies to track their emergence and spread, the observed SNPs should not be interpreted as an adaptive signature of higher viral transmissibility^27^.

Previous studies support that intra-host SARS-CoV-2 genetic diversity is transmissible and information about within-host diversity could provide further resolution for identification of geographic transmission clusters^28^ and to trace how low frequency mutations become fixed in local clusters^29^. Our analyses of intra-host diversity, however, revealed that mutations present at low-frequency (<10%) in basal strains of major Uruguayan-Brazilian clusters become highly prevalent (>80%) during subsequent transmissions, without evidence of basal samples with intermediate-frequency alternative alleles. These findings support that minor intra-host SARS-CoV-2 variants could become rapidly fixed due to stochastic events during inter-host transmissions and later perpetuated along transmission chains^30^.

In summary, our study demonstrates for the first time a recurrent viral flux across the Uruguayan-Brazilian border, with multiple introductions of the SARS-CoV-2 Brazilian lineages B.1.1.28 and B.1.1.33 into Uruguayan localities at the border region, and the potential of those viral introductions to ignite local outbreaks in Uruguay. These findings provide clear evidence that public health measures for viral control were not able to fully suppress the dissemination of SARS-CoV-2 along the Uruguayan-Brazilian border and this possess a major challenge for long-term epidemic control in Uruguay. The present work also highlights the relevance of bi-national consortiums for genomic surveillance of SARS-CoV-2 to obtain in-depth insights and better control of viral spread across highly permissive international borders.

## Data Availability

Data was uploaded to GISAID

## Acknowledgments

Thanks to the kind donors for their contribution to the project. Thanks to Carolina Banchero and Rosina Segui for beautiful figure design. Thanks to Hugo Naya for helping with statistical models and Luisa Berna for fruitful discussions. We are grateful to the Laboratory of Experimental Evolution of Viruses in charge of Gonzalo Moratorio, the Laboratory of Microbial Genomics in charge of Gregorio Iraola and the Unit of Molecular Biology in charge of Carlos Robello, all of them from the Institut Pasteur de Montevideo, for making their facilities available at any time for our experiments and sharing their experience, as well as equipment, plastic supplies and reagents. We thank Dr. Gabriela Ortiz and Dr. Paula Aguerrebere of the IAC (Instituto Asistencial Colectivo Treinta y Tres) for their willingness and hard work in contacting patients. We thank the workforce of the technical group of the Laboratory for Respiratory Viruses at LACEN / CEVS / SES-RS and the support of the Center for Scientific and Technological Development (CDCT / CEVS / SES-RS). Thanks for efforts from different groups that contribute with SARS-CoV-2 genomes to the EpiCoV GISAID initiative (Supplementary tables S9 and S10).

## Funding

Brazil: Funding support from CGLab/MoH (General Laboratories Coordination of Brazilian Ministry of Health), CVSLR/FIOCRUZ (Coordination of Health Surveillance and Reference Laboratories of Oswaldo Cruz Foundation), CNPq COVID-19 MCTI 402457/2020-0 and INOVA VPPCB-005-FIO437 20-2. Uruguay: This work was funded by the Manuel Perez Foundation that nucleated COVID19 donation fonds in the context of the project “Vigilancia epidemiológica del COVID-19 en las fronteras uruguayas y análisis de su transmisión en el interior del paí s”

## Declaration of interests

All authors declare no conflict of interest.

## Individual contributions

LS: Post processing raw sequencing data, study design, coordination of Uruguayan group, funding acquisition, verified underlying data and wrote the original draft.

DM: In charge of phylogenetic and phylogeographic analysis, verified underlying data and wrote original draft.

NR: Post processing raw sequencing data, verified underlying data, variant analysis and wrote original draft.

TFC: In charge of sequencing in Uruguay, read and corrected the manuscript.

VN: Head of diagnostic center in the Sanatorio Americano, obtaining funding, reviewed and edited the manuscript.

MA, TP, NR: Worked on diagnosis in Sanatorio Americano, read the paper.

GB: Study design, coordination between Uruguayan and Brazilian institutions, coordinated phylogeographic and phylogenetic analysis, verified underlying data and wrote the original draft.

RC: Coordination of Diagnostic centers in the Uruguayan non-metropolitan area, study design, edited and reviewed the manuscript.

FLT: Worked on diagnosis in CENUR-Norte, Uruguay, worked on Nanopore sequencing in Uruguay centers, edited and reviewed the manuscript.

MB: Worked on Nanopore sequencing in Uruguay centers and read the manuscript.

MG: Technical support at the Institut Pasteur de Montevideo, helped in writing the grant for funding, carefully edited and reviewed the manuscript.

CS: Bioinformatic analysis of sequencing data from Uruguayan centers, reviewed the manuscript.

MV, AL, MC, LM, MS: Worked on diagnosis in CENUR-Norte, Uruguay, and reviewed the manuscript.

YV: Worked on diagnosis in Laboratorio DILAVE/MGAP-INIA-UdelaR-Tacuarembó, Uruguay, and reviewed the manuscript.

CA: Worked on diagnosis in CURE-Rocha, Uruguay, and reviewed the manuscript.

CS: Worked on Nanopore sequencing at Institut Pasteur de Montevideo, Uruguay, and reviewed the manuscript.

IF: software and reviewed the manuscript

PS, JS: Worked on Ion Torrent sequencing Instituto de Investigaciones Biológicas Clemente Estable, Uruguay, and reviewed the manuscript.

RF,CM: Sequencing design, library preparation and sequencing (IonTorrent)

MJBG: Worked on logistic support and reviewed the manuscript.

IA: Worked on phylogenetic and phylogeographic analyses and reviewed the manuscript.

PCR: Coordinate the Brazilian SARS-CoV-2 sequencing team using Nanopore, Illumina, and IonTorrent platforms at the National Reference Laboratory Fiocruz, analyzed the raw sequencing data, revised and edited the manuscript.

TSG, MTMR: Perform the extraction and the SARS-CoV-2 detection by real time RT-PCR locally in the Rio Grande do Sul state, and revised the manuscript.

LGM: Responsible for the epidemiological team in the Rio Grande do Sul State, collect all the epidemiological data from Brazilian samples included in this study, and revised the manuscript.

FCM: Responsible for extraction and the SARS-CoV-2 confirmation by real time RT-PCR at the National Reference Laboratory Fiocruz, and revised the manuscript.

LA, ACM: Responsible for whole-genome SARS-CoV-2 amplification, libraries construction, and sequencing, revised the manuscript.

MMS: Financial support and revised and edited the manuscript.

## Notes

### Competing Interest Statement

The authors have declared no competing interest.

### Author Declarations

This study was approved in Uruguay by the Ethics Committees of the Sanatorio Americano (CEI-SASA). Board members were: Dr. Oscar Cluzet, Dr. Enrique Mendez, Dr. Alberto Aliaga and Mr. Richard Riera. This study was approved in Brazil by following ethics committees: FIOCRUZ-IOC Ethics Committee: 68118417.6.0000.5248, and Brazilian Ministry of Health SISGEN: A1767C3.

