## Supplementary Material for "Recurrent dissemination of SARS-CoV-2 through the Uruguayan-Brazilian border"

### SUPPLEMENTARY MATERIALS

#### Section 1. Materials and Methods

##### 1.1. SARS-CoV-2 RNA identification from samples

In the UdelaR Labs Network for SARS-CoV-2 molecular detection (CENUR Regional Norte-Salto, CURE Regional Este-Rocha, Campus Tacuarembó, Uruguay) previously described, located in Salto, Rocha and Tacuarembó cities, the viral RNA extraction was performed directly from samples (virus transport media containing the swab collected) by using the Viral RNA Isolation Kit (Liferiver Bio-Tech Corp., San Diego, USA), according to the manufacturer's instructions. The viral RNA extracted was used as a template in a OneStep RT-qPCR kit *COVID-19 RT-PCR Real TM Fast* (UdelaR, IPMON and ATGen S.R.L., Montevideo, Uruguay) to test the presence of SARS-CoV-2 RNA in the samples, according to the manufacturer's instructions. Sanatorio Americano Montevideo (SASA, Laboratory of Molecular Biology) used on the one hand, Coronavirus COVID-19 genesig® Real-Time PCR assay and on the other, GeneFinder Covid-19 Plus RealAmp Kit, according to manufacturer's instructions. In Brazil in both institutions, the viral RNA was obtained by QIAamp Viral RNA Mini kit (QIAGEN, Hilden, Germany) or automatedly Perkin-Elmer Chemagic machine/chemistry using 140 µl or 300 µl of the sample, respectively, according to the manufacturer's instructions. SARS-CoV-2 positive cases were confirmed by real-time RT-PCR assays using the SARS-CoV-2 Molecular E/RP Kit (Biomanguinhos, Rio de Janeiro, Brazil) based on the primers previously designed by[1]

##### 1.2. Reverse transcription and SARS-CoV-2 whole-genome amplification

Total RNA from positive samples was reverse transcribed using SuperScript™ IV First Strand Synthesis System (Invitrogen, Carlsbad, CA, USA), according to the manufacturer's instructions. Two multiplex PCR reactions with the primer scheme (Pool A = nine amplicons and Pool B = eight amplicons) were performed for building long (~2kb) amplicon libraries to recover SARS-CoV-2 genomes as previously described by Resende and co-workers[2], by using the Q5® High-Fidelity DNA Polymerase (New England Biolabs, Ipswich, MA, USA), according to the manufacturer's instructions. When SARS-CoV-2 genome amplification was unsuccessful with the long 2kb amplicons strategy, a second strategy was used. In this case SARS-CoV-2 genomes were recovered by building short (~400pb) amplicon libraries as previously described by Quick and co-workers[3], using the hCoV-19 primer scheme V3 (which can be obtained under https://github.com/artic-network/primer-schemes/tree/master/nCoV-2019/V3). Both protocols (long and short amplicons libraries) are based on the amplicon tiling strategy described previously by Quick and co-workers[4]. Both pools were mixed and the amplicons were purified using Agencourt AMPure XP beads (Beckman Coulter™, Brea, CA, USA) and a quality control was performed to measure the quantity of DNA using the Qubit™ dsDNA BR Assay Kit (Invitrogen) and Qubit Fluorometric Quantification.

#### 1.3. SARS-CoV-2 whole-genome sequencing

Different sequencing methods were used to produce the genomes for this study, such as Illumina, Nanopore and Ion Torrent. ONT libraries were prepared by using the Ligation Sequencing Kit (SQK-LSK109) and Native Barcoding Expansion (EXP-NBD104 and EXP-NBD114), both from ONT (Oxford Nanopore Technologies, United Kingdom). The NEBNext Ultra II End Repair/dA-Tailing Module and NEBNext Ultra II Ligation Module were used to ligate barcodes and sequence adapters to each sample (New England Biolabs, Ipswich, MA, USA), according to the manufacturer's instructions. Negative controls using H2O as template (no RNA) in the RT step were added as amplification controls and were kept in the rest of the process as sequencing controls. We used MinION sequencing platforms (Oxford Nanopore Technologies Ltd., Oxford, UK) as described in detail elsewhere[5]. Illumina short reads DNA libraries were generated from the pooled amplicons using Nextera XT DNA Sample Preparation Kit (Illumina, San Diego, CA, USA) according to the manufacturer specifications. Library sizes were evaluated using a 4200 TapeStation System (Agilent, Santa Clara, USA). Samples were then sequenced (pair-end) (Micro V2, 300 cycles) on a MiSeq equipment (Illumina, San Diego, USA) in around 18 hours. Ion Torrent: 100 ng of each purified PCR products were used to produce Ion Torrent™ compatible sequencing libraries, using NEBNext® Fast DNA Library Prep Set for Ion Torrent™ (#E6270L, New England Biolabs, Inc) and Ion Xpress™ Barcode Adapters (#4474517, Ion Torrent™) following manufacturer instructions. Following Qubit™ Fluorometer quantification with Qubit™ dsDNA HS Assay Kit, libraries were diluted and combined in a equimolar fashion to obtain 25 µl of 50 pM the input for the sequencing template preparation. Template preparation and Ion 530™ Chip loading were conducted with the Ion Chef™ Instrument, with the Ion 510™ & Ion 520™ & Ion 530™ Kit – Chef (#A34019, Ion Torrent™), following manufacturer instructions. Sequencing was performed in the Ion GeneStudio™ S5 System™, using Ion 510™ & Ion 520™ & Ion 530™ Kit – Chef (#A34019, Ion Torrent™), following manufacturer instructions.

#### 1.4. SARS-CoV-2 whole-genome consensus sequences

Each of the sequencing platforms used generated fastq files analyzed by different methods to obtain final consensus sequence.

The raw data from the MinION was analyzed with the pipeline developed by the COVID-19 Genomics UK consortium[6], which is based on the Artic Network bioinformatic pipeline[7]. The pipeline was embedded within Nextflow as in[8] with minor modifications to fit local infrastructure[9].

Within that pipeline different combinations of parameters were used. For the demultiplexing step with Guppy v3.6.0 (https://community.nanoporetech.com) we asked for both barcodes to be present (at both ends with min_score 50 and min_score_rear_override 40). Alternatively, we requested one barcode with a higher score (min_score 60). Careful inspection of results indicated that relaxing Guppy parameters decreased N content in the final sequence without introducing any variants.

Ion Torrent reads were mapped to the reference genome using Bowtie 2[10] allowing local alignments. Finally, bcftools was used for SNP calling (mpileup), SNP filtering (minimum quality of 20 and filtering adjacent indels within 5 bases) and to build the consensus sequences (consensus function). Positions of interest (about 20 bases around synapomorphic sites) were manually inspected to resolve undetermined bases.

In Brazilian Institutions, demultiplexed fastq files generated by Illumina or IonTorrent sequencing were used as the input for the analysis. Reads were trimmed based on quality scores with a cutoff of Q30, in order to remove low quality regions and adapter sequences were filtered. Following standard pre-processing steps, reads were mapped to the hCoV-19/Wuhan/Hu-1/2019 strain (GISAID accession number EPI_ISL_402125). Duplicate reads were removed from the alignment and the consensus sequence called at a threshold of 10x. The entire workflow was carried out in CLC Genomics Workbench software version 20.0 (https://digitalinsights.qiagen.com).

#### 1.5 Maximum likelihood phylogenetic analyses

All full-length B.1.1.28 (n = 275) and B.1.1.33 (n = 492) SARS-CoV-2 genomes available on GISAID (https://www.gisaid.org/) as of October 19 2020, were downloaded and aligned with Uruguayan and Brazilian sequences of the same lineages generated in this study using MAFFT v7.467[11]. ML phylogenetic trees were constructed under the GTR+F+I+G4 nucleotide substitution model selected by the in-built Smart Model Selection (SMS) option[12] and visualized using FigTree v1.4.4 (http://tree.bio.ed.ac.uk/software/figtree/).

#### 1.6 Bayesian phylogeographic analyses

Temporal signal was assessed by performing a regression analysis of the root-to-tip divergence against sampling time using TempEst[13]. Time-scaled Bayesian trees were estimated using the non-parametric Bayesian skyline (BSKL) model as the coalescent tree prior[14], under the optimal model of nucleotide substitution determined for each data set by the SMS option and applying a strict molecular clock model with a fixed substitution rate ($8 \times 10^{-4}$ substitutions/site/year) based on previous estimates[15,16]. Three MCMC chains were run for 100 and 200 million generations for the B.1.1.28 and B.1.1.33 datasets, respectively, and then combined to ensure stationarity and good mixing. Convergence (Effective Sample Size > 200) in parameter estimates was assessed using TRACER v1.7[17]. The maximum clade credibility (MCC) trees were summarized with TreeAnnotator v1.10 and visualized using FigTree v1.4.4.

#### 1.7 Within host diversity

Given that for some samples read coverage was low at the identified synapomorphic sites, base frequencies were obtained straight from the adapter trimmed bam files (Tables S4-S7). Only those observations where the genome position was covered by at least 100 reads were kept for further analysis. As samples were obtained by three different sequencing technologies (data for 108, 108 and 494 synapomorphic sites was obtained using Illumina, Ion Torrent and ONT, respectively) and sequencing biases exist due to different error profiles, a Shannon Entropy[18] value (H) was estimated per observation based on the four allele frequencies. As diagnostics plots evidence an effect due to the sequencing technology (Figures S1A-B), we determined a linear model to estimate the contributions of sequencing technology, sample and synapomorphic site, to the observed A, C, G, T frequencies (summarized as H): H exp ¼ ~ SEQ + SAMPLE + MUTATION. We used H exp ¼ to better fit a normal distribution (Figure S1C). This was implemented using the lm function of the R environment[19]. Residuals of the model are shown in Figure S1D and coefficients for the covariables are shown in Table S8.

#### Section 2. Supplementary tables

**Table S1:** Detailed information of each genome generated for this study by the different sequencing centers. Travel "yes" means that person had recently traveled or was in contact with a recent traveler. Superscript B means that the demultiplexing step was done requiring both barcodes at both ends with min_score above 50 and 40, respectively. Superscript R corresponds to the demultiplexing step without requiring the presence of both barcodes but using min_score above 60.

| Diagnostic Center | Sequencing Center | Accession ID | CollectionSite | Sequencing Method | Protocol | Travel |
|---|---|---|---|---|---|---|
| SASA | Institut Pasteur de Montevideo | ART-M107 | Artigas | MinION | V3 short amplicons$^{R}$ | |
| CENUR Regional Norte | Institut Pasteur de Montevideo | ART-M146 | Artigas | MinION | V3 short amplicons$^{R}$ | yes |
| CENUR Regional Norte | Institut Pasteur de Montevideo | BUN-M151 | Bella Unión | MinION | long amplicons$^{R}$ | |
| CENUR Regional Norte | Institut Pasteur de Montevideo | BUN-M152 | Bella Unión | MinION | long amplicons$^{R}$ | |
| CENUR Regional Norte | Institut Pasteur de Montevideo | BUN-M163 | Bella Unión | MinION | V3 short amplicons$^{R}$ | |
| SASA | Institut Pasteur de Montevideo | CEL-M116 | Río Branco | MinION | long amplicons$^{R}$ | |
| CURE, Regional Este | Institut Pasteur de Montevideo | CHY-M100 | Chuy | MinION | V3 short amplicons$^{R}$ | |
| CENUR Regional Norte | IIBCE | RIV-M10 | Rivera | Ion Torrent | V3 short amplicons | |
| CENUR Regional Norte | IIBCE | RIV-M11 | Rivera | Ion Torrent | V3 short amplicons | |
| SASA | Institut Pasteur de Montevideo | RIV-M119 | Rivera | MinION | long amplicons$^{R}$ | |
| CENUR Regional Norte | IIBCE | RIV-M12 | Rivera | Ion Torrent | V3 short amplicons | |
| CENUR Regional Norte | IIBCE | RIV-M13 | Rivera | Ion Torrent | V3 short amplicons | |
| CENUR Regional Norte | IIBCE | RIV-M14 | Rivera | Ion Torrent | V3 short amplicons | |
| CENUR | IIBCE | RIV-M15 | Rivera | Ion Torrent | V3 short | |

| | | | | | |
|---|---|---|---|---|---|
| Regional Norte | | | | | amplicons |
| CENUR Regional Norte | Institut Pasteur de Montevideo | RIV-M154 | Rivera | MinION | long amplicons[R] |
| CENUR Regional Norte | Institut Pasteur de Montevideo | RIV-M155 | Rivera | MinION | V3 short amplicons[R] |
| Campus Tacuarembó | Institut Pasteur de Montevideo | RIV-M170 | Rivera | MinION | long amplicons[R] |
| SASA | Institut Pasteur de Montevideo | RIV-M40 | Rivera | MinION | long amplicons[B] |
| SASA | Institut Pasteur de Montevideo | RIV-M43 | Rivera | MinION | V3 short amplicons[B] |
| CENUR Regional Norte | IIBCE | RIV-M5 | Rivera | Ion Torrent | V3 short amplicons |
| Campus Tacuarembó | Institut Pasteur de Montevideo | RIV-M72 | Rivera | MinION | V3 short amplicons[B] |
| Campus Tacuarembó | Institut Pasteur de Montevideo | RIV-M73 | Rivera | MinION | V3 short amplicons[B] |
| CENUR Regional Norte | Institut Pasteur de Montevideo | RIV-M79 | Rivera | MinION | V3 short amplicons[B] |
| CENUR Regional Norte | IIBCE | RIV-M8 | Rivera | Ion Torrent | V3 short amplicons |
| CENUR Regional Norte | IIBCE | RIV-M9 | Rivera | Ion Torrent | V3 short amplicons |
| CENUR Regional Norte | Institut Pasteur de Montevideo | ART-M157 | Artigas | MinION | V3 short amplicons[R] |
| SASA | Institut Pasteur de Montevideo | TYT-M130 | Treinta y Tres | MinION | V3 short amplicons[R] |
| SASA | Institut Pasteur de Montevideo | TYT-M132 | Treinta y Tres | MinION | V3 short amplicons[R] |
| SASA | Institut Pasteur de Montevideo | TYT-M133 | Treinta y Tres | MinION | long amplicons[R] |
| SASA | Institut Pasteur de Montevideo | TYT-M135 | Treinta y Tres | MinION | long amplicons[R] |

| | | | | | | |
|---|---|---|---|---|---|---|
| SASA | Institut Pasteur de Montevideo | TYT-M136 | Treinta y Tres | MinION | long amplicons[R] | |
| SASA | Institut Pasteur de Montevideo | TYT-M49 | Treinta y Tres | MinION | long amplicons[B] | yes |
| SASA | Institut Pasteur de Montevideo | TYT-M52 | Treinta y Tres | MinION | V3 short amplicons[R] | |
| SASA | Institut Pasteur de Montevideo | TYT-M54 | Treinta y Tres | MinION | V3 short amplicons[R] | |
| SASA | Institut Pasteur de Montevideo | TYT-M57 | Treinta y Tres | MinION | long amplicons[B] | |
| SASA | Institut Pasteur de Montevideo | TYT-M60 | Treinta y Tres | MinION | V3 short amplicons[R] | |
| SASA | Institut Pasteur de Montevideo | TYT-M62 | Treinta y Tres | MinION | V3 short amplicons[R] | |
| SASA | Institut Pasteur de Montevideo | TYT-M63 | Treinta y Tres | MinION | V3 short amplicons[R] | |
| SASA | Institut Pasteur de Montevideo | TYT-M64 | Treinta y Tres | MinION | long amplicons[B] | |
| SASA | Institut Pasteur de Montevideo | TYT-M65 | Treinta y Tres | MinION | V3 short amplicons[R] | |
| SASA | Institut Pasteur de Montevideo | TYT-M66 | Treinta y Tres | MinION | V3 short amplicons[R] | |
| SASA | Institut Pasteur de Montevideo | TYT-M67 | Treinta y Tres | MinION | V3 short amplicons[R] | |
| SASA | Institut Pasteur de Montevideo | TYT-M68 | Treinta y Tres | MinION | V3 short amplicons[R] | |
| SASA | Institut Pasteur de Montevideo | TYT-M69 | Treinta y Tres | MinION | V3 short amplicons[R] | |
| SASA | Institut Pasteur de Montevideo | TYT-M70 | Treinta y Tres | MinION | V3 short amplicons[R] | |
| SASA | Institut Pasteur de Montevideo | TYT-M71 | Treinta y Tres | MinION | V3 short amplicons[R] | |
| SASA | Institut Pasteur de Montevideo | TYT-M85 | Treinta y Tres | MinION | V3 short amplicons[B] | |
| SASA | Institut Pasteur de Montevideo | TYT-M87 | Treinta y Tres | MinION | V3 short amplicons[B] | |

| | | | | | |
|---|---|---|---|---|---|
| SASA | Institut Pasteur de Montevideo | TYT-M91 | Treinta y Tres | MinION | long amplicons[B] |
| SASA | Institut Pasteur de Montevideo | TYT-M93 | Treinta y Tres | MinION | V3 short amplicons[B] |
| SASA | Institut Pasteur de Montevideo | TYT-M94 | Treinta y Tres | MinION | V3 short amplicons[B] |
| SASA | Institut Pasteur de Montevideo | TYT-M95 | Treinta y Tres | MinION | V3 short amplicons[B] |
| SASA | Institut Pasteur de Montevideo | TYT-M97 | Treinta y Tres | MinION | long amplicons[B] |
| SASA | Institut Pasteur de Montevideo | TYT-M98 | Treinta y Tres | MinION | V3 short amplicons[B] |
| LACEN/RS | LRN, IOC, Fiocruz | hCoV-19/Brazil/RS-15270/2020 | PASSO FUNDO | Illumina Miseq | long amplicons |
| LACEN/RS | LRN, IOC, Fiocruz | hCoV-19/Brazil/RS-15273/2020 | CACHOEIRA DO SUL | Illumina Miseq | long amplicons |
| LACEN/RS | LRN, IOC, Fiocruz | hCoV-19/Brazil/RS-15279/2020 | NOVA BRESCIA | Illumina Miseq | long amplicons |
| LACEN/RS | LRN, IOC, Fiocruz | hCoV-19/Brazil/RS-15283/2020 | PORTO ALEGRE | Illumina Miseq | long amplicons |
| LACEN/RS | LRN, IOC, Fiocruz | hCoV-19/Brazil/RS-15286/2020 | SAO BORJA | Illumina Miseq | long amplicons |
| LACEN/RS | LRN, IOC, Fiocruz | hCoV-19/Brazil/RS-2525/2020 | PORTO ALEGRE | Illumina Miseq | long amplicons |
| LACEN/RS | LRN, IOC, Fiocruz | hCoV-19/Brazil/RS-2528/2020 | ERECHIM | Illumina Miseq | long amplicons |
| LACEN/RS | LRN, IOC, Fiocruz | hCoV-19/Brazil/RS-2529/2020 | SANTANA DO LIVRAMENTO | Illumina Miseq | long amplicons |
| LACEN/RS | LRN, IOC, Fiocruz | hCoV-19/Brazil/RS-2533/2020 | TORRES | Illumina Miseq | long amplicons |
| LACEN/RS | LRN, IOC, Fiocruz | hCoV-19/Brazil/RS-2539/2020 | SANTA MARIA | Illumina Miseq | long amplicons |
| LACEN/RS | LRN, IOC, Fiocruz | hCoV-19/Brazil/RS-2544/2020 | TORRES | Illumina Miseq | long amplicons |
| LACEN/RS | LRN, IOC, | hCoV- | PORTO | Illumina | long |

| | | | | | |
|---|---|---|---|---|---|
| | Fiocruz | 19/Brazil/RS-2546/2020 | ALEGRE | Miseq | amplicons |
| LACEN/RS | LRN, IOC, Fiocruz | hCoV-19/Brazil/RS-2549/2020 | MARAU | Illumina Miseq | long amplicons |
| LACEN/RS | LRN, IOC, Fiocruz | hCoV-19/Brazil/RS-2550/2020 | SÃO LEOPOLDO | Illumina Miseq | long amplicons |
| LACEN/RS | LRN, IOC, Fiocruz | hCoV-19/Brazil/RS-2553/2020 | SÃO DOMINGOS DO SUL | Illumina Miseq | long amplicons |
| LACEN/RS | LRN, IOC, Fiocruz | hCoV-19/Brazil/RS-2554/2020 | PORTO ALEGRE | Illumina Miseq | long amplicons |
| LACEN/RS | LRN, IOC, Fiocruz | hCoV-19/Brazil/RS-2556/2020 | FARROUPILHA | Illumina Miseq | long amplicons |
| LACEN/RS | LRN, IOC, Fiocruz | hCoV-19/Brazil/RS-2564/2020 | PORTO ALEGRE | Illumina Miseq | long amplicons |
| LACEN/RS | LRN, IOC, Fiocruz | hCoV-19/Brazil/RS-2565/2020 | PORTO ALEGRE | Illumina Miseq | long amplicons |
| LACEN/RS | LRN, IOC, Fiocruz | hCoV-19/Brazil/RS-2567/2020 | CIDREIRA | Illumina Miseq | long amplicons |
| LACEN/RS | LRN, IOC, Fiocruz | hCoV-19/Brazil/RS-6169/2020 | PASSO FUNDO | Illumina Miseq | long amplicons |
| LACEN/RS | LRN, IOC, Fiocruz | hCoV-19/Brazil/RS-6177/2020 | CARLOS BARBOSA | Illumina Miseq | long amplicons |
| LACEN/RS | LRN, IOC, Fiocruz | hCoV-19/Brazil/RS-6179/2020 | SANTA ROSA | Illumina Miseq | long amplicons |
| LACEN/RS | LRN, IOC, Fiocruz | hCoV-19/Brazil/RS-6180/2020 | VENÂNCIO AIRES | Illumina Miseq | long amplicons |
| LACEN/RS | LRN, IOC, Fiocruz | hCoV-19/Brazil/RS-6183/2020 | SALDANHA MARINHO | Illumina Miseq | long amplicons |
| LACEN/RS | LRN, IOC, Fiocruz | hCoV-19/Brazil/RS-6184/2020 | ITAQUI | Illumina Miseq | long amplicons |
| LACEN/RS | LRN, IOC, Fiocruz | hCoV-19/Brazil/RS-6187/2020 | VILA MARIA | Illumina Miseq | long amplicons |
| LACEN/RS | LRN, IOC, | hCoV- | CAXIAS DO | Illumina | long |

| | | | | | |
|---|---|---|---|---|---|
| | Fiocruz | 19/Brazil/RS-6188/2020 | SUL | Miseq | amplicons |
| LACEN/RS | LRN, IOC, Fiocruz | hCoV-19/Brazil/RS-6189/2020 | CAXIAS DO SUL | Illumina Miseq | long amplicons |
| LACEN/RS | LRN, IOC, Fiocruz | hCoV-19/Brazil/RS-6190/2020 | ARROIO DO MEIO | Illumina Miseq | long amplicons |
| LACEN/RS | LRN, IOC, Fiocruz | hCoV-19/Brazil/RS-6192/2020 | SERAFINA CORREA | Illumina Miseq | long amplicons |
| LACEN/RS | LRN, IOC, Fiocruz | hCoV-19/Brazil/RS-6195/2020 | NOVA ARAÇA | Illumina Miseq | long amplicons |
| LACEN/RS | LRN, IOC, Fiocruz | hCoV-19/Brazil/RS-6196/2020 | PASSO FUNDO | Illumina Miseq | long amplicons |
| LACEN/RS | LRN, IOC, Fiocruz | hCoV-19/Brazil/RS-6197/2020 | NÃO ME TOQUE | Illumina Miseq | long amplicons |
| LACEN/RS | LRN, IOC, Fiocruz | hCoV-19/Brazil/RS-6198/2020 | SERAFINA CORREA | Illumina Miseq | long amplicons |
| LACEN/RS | LRN, IOC, Fiocruz | hCoV-19/Brazil/RS-6203/2020 | QUARAÍ | Illumina Miseq | long amplicons |
| LACEN/RS | LRN, IOC, Fiocruz | hCoV-19/Brazil/RS-6205/2020 | SANTANA DO LIVRAMENTO | Illumina Miseq | long amplicons |
| LACEN/RS | LRN, IOC, Fiocruz | hCoV-19/Brazil/RS-6208/2020 | FREDERICO WESTPHALEN | Illumina Miseq | long amplicons |
| LACEN/RS | LRN, IOC, Fiocruz | hCoV-19/Brazil/RS-6213/2020 | PASSO FUNDO | Illumina Miseq | long amplicons |
| LACEN/RS | LRN, IOC, Fiocruz | hCoV-19/Brazil/RS-6215/2020 | PASSO FUNDO | Illumina Miseq | long amplicons |
| LACEN/RS | LRN, IOC, Fiocruz | hCoV-19/Brazil/RS-6218/2020 | VAIMAO | Illumina Miseq | long amplicons |
| LACEN/RS | LRN, IOC, Fiocruz | hCoV-19/Brazil/RS-6219/2020 | PORTO ALEGRE | Illumina Miseq | long amplicons |
| LACEN/RS | LRN, IOC, Fiocruz | hCoV-19/Brazil/RS-6220/2020 | PORTO ALEGRE | Illumina Miseq | long amplicons |
| LACEN/RS | LRN, IOC, Fiocruz | hCoV-19/Brazil/RS-6222/2020 | FLORES DA CUNHA | Illumina Miseq | long amplicons |
| LACEN/RS | LRN, IOC, Fiocruz | hCoV-19/Brazil/RS- | VENANCIO AIRES | Illumina Miseq | long amplicons |

| | | | | | |
|---|---|---|---|---|---|
| | | 6227/2020 | | | |
| LACEN/RS | LRN, IOC, Fiocruz | hCoV-19/Brazil/RS-6228/2020 | LAJEADO | Illumina Miseq | long amplicons |
| LACEN/RS | LRN, IOC, Fiocruz | hCoV-19/Brazil/RS-6231/2020 | CAÇAPAVA DO SUL | Illumina Miseq | long amplicons |
| LACEN/RS | LRN, IOC, Fiocruz | hCoV-19/Brazil/RS-6232/2020 | BAGE | Illumina Miseq | long amplicons |
| LACEN/RS | LRN, IOC, Fiocruz | hCoV-19/Brazil/RS-6240/2020 | SANTO ANGELO | Illumina Miseq | long amplicons |
| LACEN/RS | LRN, IOC, Fiocruz | hCoV-19/Brazil/RS-6241/2020 | GIRUA | Illumina Miseq | long amplicons |
| LACEN/RS | LRN, IOC, Fiocruz | hCoV-19/Brazil/RS-6242/2020 | CRUZ ALTA | Illumina Miseq | long amplicons |
| LACEN/RS | LRN, IOC, Fiocruz | hCoV-19/Brazil/RS-6226/2020 | BARRA DO RIBEIRO | IonTorrent | long amplicons |
| LACEN/RS | LRN, IOC, Fiocruz | hCoV-19/Brazil/RS-6243/2020 | IGREJINHA | IonTorrent | long amplicons |
| LACEN/RS | LRN, IOC, Fiocruz | hCoV-19/Brazil/RS-15274/2020 | PORTO ALEGRE | Nanopore MinION | long amplicons |
| LACEN/RS | LRN, IOC, Fiocruz | hCoV-19/Brazil/RS-15275/2020 | ARROIO GRANDE | Nanopore MinION | long amplicons |
| LACEN/RS | LRN, IOC, Fiocruz | hCoV-19/Brazil/RS-15276/2020 | BOM RETIRO DO SUL | Nanopore MinION | long amplicons |
| LACEN/RS | LRN, IOC, Fiocruz | hCoV-19/Brazil/RS-15278/2020 | TAPEJARA | Nanopore MinION | long amplicons |
| LACEN/RS | LRN, IOC, Fiocruz | hCoV-19/Brazil/RS-15280/2020 | PORTO ALEGRE | Nanopore MinION | long amplicons |
| LACEN/RS | LRN, IOC, Fiocruz | hCoV-19/Brazil/RS-15281/2020 | FARROUPILHA | Nanopore MinION | long amplicons |
| LACEN/RS | LRN, IOC, Fiocruz | hCoV-19/Brazil/RS-15282/2020 | CAXIAS DO SUL | Nanopore MinION | long amplicons |
| LACEN/RS | LRN, IOC, Fiocruz | hCoV-19/Brazil/RS-15284/2020 | IJUI | Nanopore MinION | long amplicons |
| LACEN/RS | LRN, IOC, Fiocruz | hCoV-19/Brazil/RS-15285/2020 | VENANCIO AIRES | Nanopore MinION | long amplicons |
| LACEN/RS | LRN, IOC, | hCoV- | SANTA ROSA | Nanopore | long |

| | | | | | |
|---|---|---|---|---|---|
| | Fiocruz | 19/Brazil/RS-15287/2020 | | MinION | amplicons |
| LACEN/RS | LRN, IOC, Fiocruz | hCoV-19/Brazil/RS-15288/2020 | NAO-ME-TOQUE | Nanopore MinION | long amplicons |
| LACEN/RS | LRN, IOC, Fiocruz | hCoV-19/Brazil/RS-15289/2020 | PORTO ALEGRE | Nanopore MinION | long amplicons |
| LACEN/RS | LRN, IOC, Fiocruz | hCoV-19/Brazil/RS-15290/2020 | OSORIO | Nanopore MinION | long amplicons |
| LACEN/RS | LRN, IOC, Fiocruz | hCoV-19/Brazil/RS-15291/2020 | TRES PASSOS | Nanopore MinION | long amplicons |
| LACEN/RS | LRN, IOC, Fiocruz | hCoV-19/Brazil/RS-15292/2020 | PORTO ALEGRE | Nanopore MinION | long amplicons |

#### Table S2. Accession IDs Uruguayan Genomes

| Accession ID |
|---|
| EPI_ISL_747615 |
| EPI_ISL_749238 |
| EPI_ISL_748667 |
| EPI_ISL_748143 |
| EPI_ISL_748142 |
| EPI_ISL_749036 |
| EPI_ISL_748141 |
| EPI_ISL_748140 |
| EPI_ISL_749152 |
| EPI_ISL_749153 |
| EPI_ISL_748145 |
| EPI_ISL_749154 |
| EPI_ISL_748144 |
| EPI_ISL_749155 |
| EPI_ISL_749474 |
| EPI_ISL_749150 |
| EPI_ISL_749151 |
| EPI_ISL_750108 |
| EPI_ISL_750430 |
| EPI_ISL_750820 |
| EPI_ISL_749906 |
| EPI_ISL_748139 |
| EPI_ISL_748138 |
| EPI_ISL_749706 |
| EPI_ISL_749149 |

| |
|---|
| EPI_ISL_749148 |
| EPI_ISL_750167 |
| EPI_ISL_750165 |
| EPI_ISL_750166 |
| EPI_ISL_750163 |
| EPI_ISL_750164 |
| EPI_ISL_751011 |
| EPI_ISL_750161 |
| EPI_ISL_750162 |
| EPI_ISL_750169 |
| EPI_ISL_750170 |
| EPI_ISL_750171 |
| EPI_ISL_750178 |
| EPI_ISL_750179 |
| EPI_ISL_750256 |
| EPI_ISL_750176 |
| EPI_ISL_750177 |
| EPI_ISL_750174 |
| EPI_ISL_750175 |
| EPI_ISL_750172 |
| EPI_ISL_751184 |
| EPI_ISL_750173 |
| EPI_ISL_751186 |
| EPI_ISL_751187 |
| EPI_ISL_751185 |
| EPI_ISL_751190 |
| EPI_ISL_751201 |
| EPI_ISL_751188 |
| EPI_ISL_751189 |

**Table S3.** SARS-CoV-2 lineage-defining SNPs. Lineage-defining SNPs (mutations), that were sequentially fixed during evolution and dissemination of each SARS-CoV-2 Brazilian-Uruguayan clades here identified, $T_{MRCA}$ and samples that compose each clade.

| Clade | Subclade | Subclade | Subclade | Accession ID |
|---|---|---|---|---|
| | | | | D-NVRL-72IRL12139 |
| | | | | RJ-1901 |
| | | | | RJ-2000 |
| | | | | RJ-2007 |
| | | | | RJ-2091 |
| | | | | RJ-2342 |

| | | | | |
|---|---|---|---|---|
| | | | | SP-259 |
| | | | | SP-263 |
| | | | | SP-315 |
| | | | | SP-564 |
| | | | | SP-L7-CD72 |
| | | | | TyT-M70 |
| | | | | TyT-M132 |
| | | | | TyT-M133 |
| | | | | TyT-M135 |
| | | | | TyT-M136 |
| | | | | TyT-M49 |
| | | | | TyT-M52 |
| | | | | TyT-M54 |
| | | | | TyT-M57 |
| **BR-UY-$I_{33}$** | | | | TyT-M60 |
| **S:22720A>G** | | | | TyT-M62 |
| **Mar 12** | **TT-$I_{33}$ + SP** | | | TyT-M63 |
| **(Mar 09- Mar 21)** | **ORF1ab:8293C>T** | | | TyT-M64 |
| | **Mar 17** | **TT-$I_{33}$** | | TyT-M65 |
| | **(Mar 14-Mar 25)** | **ORF1ab:5089A>G**<br>**N:28775C>T** | | TyT-M66 |
| | | **May 26** | **TT-$I_{33}$ less one** | TyT-M67 |
| | | **(May 12-Jun 07)** | **ORF1ab:15108C>T** | TyT-M68 |
| | | | **May 30** | TyT-M69 |
| | | | **(May 22-Jun 06)** | TyT-M130 |
| | | | | TyT-M71 |
| | | | | TyT-M85 |
| | | | | TyT-M87 |
| | | | | TyT-M91 |
| | | | | TyT-M93 |
| | | | | TyT-M94 |
| | | | | TyT-M95 |
| | | | | TyT-M97 |
| | | | | TyT-M98 |
| | | | | RS-6177 |
| | | | | RS-15280 |
| | | | | RS-6222 |
| | | | | RS-15273 |
| | | | | RS-6188 |
| | | | | RS-6203 |

| | | | | |
|---|---|---|---|---|
| | | | | RS-6208 |
| | | | | RS-6231 |
| | | | RI-II$_{33}$ | RIV-M119 |
| | | | 5’UTR:189G>T<br>ORF1ab:12025C>T<br>S:23311G>T | RIV-M154 |
| | | | July 10<br>(Jun 27-Jul 18) | RIV-M155 |
| BR-UY-II$_{33}$ | | | | RIV-M10 |
| ORF8:28093C>T | | | | RIV-M11 |
| Mar 29 | RI-I$_{33}$ +RS | | | RIV-M12 |
| (Mar 12-Apr 13) | ORF1ab:18252C>T<br>ORF1ab:2276A>G | | | RIV-M13 |
| | Apr 16 | RI-I$_{33}$ | | RIV-M14 |
| | (Apr 05-Apr 30) | S:24872G>T | | RIV-M15 |
| | | May 2 | | RIV-M40 |
| | | (Apr 27-May 04) | | RIV-M43 |
| | | | | RIV-M5 |
| | | | | RIV-M72 |
| | | | | RIV-M73 |
| | | | | RIV-M79 |
| | | | | RIV-M8 |
| | | | | RIV-M9 |
| | | | | UY-650 |
| | | | | UY-651 |
| | | | | UY-652 |
| | | | | UY-667 |
| | | | | UY-957 |
| | | | | SP-275 |
| | AR-I$_{28}$<br>ORF1ab:1471C>T,<br>1519C>T, 5995A>G,<br>9605T>C, 15606T>C | | | ART-M157 |
| BR-UY-I$_{28}$ | S:23416A>T<br>ORF3a:26159T>C<br>N:28508A>T, 29367C>T<br>3’UTR:29737G>T | | | BUN-M151 |
| | Jul 02 | | | BUN-M152 |
| | (Jun 17-Jul 16) | | | BUN-M163 |
| | | | | RS-L19-CD390 |
| | | | | RS-15276 |
| | | | | RS-6242 |

| | | |
|---|---|---|
| **BR-RS-I33** | | RS-6228 |
| **ORF1ab:15654C>T** | | RS-6179 |
| **Mar 23** | | RS-6190 |
| **(Mar 14- Mar 31)** | | RS-6197 |
| | | RS-6198 |
| | | RS-6241 |
| | | RS-15287 |
| | | RS-6187 |
| | | RS-6227 |
| | | RS-6180 |

**Table S4.** Within-host diversity BR-UY-II33. The table shows the observed allele count for each synapomorphic site identified in clade BR-UY-II33

| **CLADE BR-UY-II33** | | | | | | | | | | | | |
|---|---|---|---|---|---|---|---|---|---|---|---|---|
| | | | **1. 189G>T (5'-UTR)** | | | | | **2. 2276A>G (ORF1ab: I671V)** | | | | |
| Lineage | Outbreak | Sample | A | C | G | T | num.reads | A | C | G | T | num.reads |
| B.1.1.33 | Rivera I | M5 | 1 | 0 | 21564 | 0 | 21565 | 91 | 0 | 8177 | 6 | 8274 |
| B.1.1.33 | Rivera I | M8 | 1 | 1 | 15403 | 0 | 15405 | 226 | 0 | 545 | 0 | 771 |
| B.1.1.33 | Rivera I | M9 | 2 | 1 | 14404 | 0 | 14407 | 24 | 0 | 1746 | 1 | 1771 |
| B.1.1.33 | Rivera I | M10 | 0 | 3 | 11145 | 1 | 11149 | 47 | 0 | 3693 | 5 | 3745 |
| B.1.1.33 | Rivera I | M11 | 0 | 0 | 17862 | 0 | 17862 | 64 | 0 | 4760 | 10 | 4834 |
| B.1.1.33 | Rivera I | M12 | 1 | 1 | 14172 | 0 | 14174 | 35 | 0 | 3019 | 4 | 3058 |
| B.1.1.33 | Rivera I | M13 | 0 | 3 | 12904 | 0 | 12907 | 59 | 0 | 4083 | 5 | 4147 |
| B.1.1.33 | Rivera I | M14 | 3 | 1 | 12670 | 0 | 12674 | 66 | 0 | 5239 | 8 | 5313 |
| B.1.1.33 | Rivera I | M15 | 0 | 0 | 19821 | 0 | 19821 | 53 | 0 | 4705 | 12 | 4770 |
| B.1.1.33 | Rivera I | M40 | 3 | 0 | 195 | 0 | 198 | 45 | 2 | 882 | 0 | 929 |
| B.1.1.33 | Rivera I | M43 | 12 | 1 | 969 | 4 | 986 | 34 | 0 | 630 | 1 | 665 |
| B.1.1.33 | Rivera I | M72 | 7 | 0 | 984 | 1 | 992 | 35 | 2 | 628 | 2 | 667 |
| B.1.1.33 | Rivera I | M73 | 7 | 2 | 979 | 0 | 988 | 0 | 0 | 3 | 0 | 3 |
| B.1.1.33 | Rivera I | M79 | 8 | 1 | 978 | 2 | 989 | 9 | 0 | 133 | 0 | 142 |
| B.1.1.33 | Rivera II | M119 | 0 | 1 | 3 | 158 | 162 | 716 | 0 | 5 | 1 | 722 |
| B.1.1.33 | Rivera II | M154 | 0 | 2 | 4 | 135 | 141 | 562 | 0 | 9 | 2 | 573 |
| B.1.1.33 | Rivera II | M155 | 2 | 9 | 31 | 946 | 988 | 132 | 0 | 2 | 0 | 134 |
| B.1.1.33 | | RS-15273 | 0 | 1 | 1267 | 2 | 1270 | 1 | 0 | 1895 | 4 | 1900 |
| B.1.1.33 | | RS-15280 | 2 | 0 | 318 | 0 | 320 | 476 | 1 | 12 | 0 | 489 |
| B.1.1.33 | | RS-6177 | 2 | 1 | 960 | 2 | 965 | 3539 | 14 | 5 | 11 | 3569 |
| B.1.1.33 | | RS-6188 | 0 | 0 | 116 | 0 | 116 | 0 | 0 | 2251 | 2 | 2253 |
| B.1.1.33 | | RS-6203 | 0 | 0 | 259 | 0 | 259 | 1 | 5 | 2954 | 6 | 2966 |

| | | | | | | | | | | | | |
|---|---|---|---|---|---|---|---|---|---|---|---|---|
| B.1.1.33 | | RS-6208 | 1 | 0 | 144 | 1 | 146 | 136 | 1 | 945 | 1 | 1083 |
| B.1.1.33 | | RS-6222 | 0 | 1 | 305 | 1 | 307 | 1 | 1 | 1089 | 0 | 1091 |
| B.1.1.33 | | RS-6231 | 0 | 0 | 111 | 0 | 111 | 0 | 1 | 687 | 1 | 689 |
| | | | **3. 12025C>T (ORF1ab: S3920S)** | | | | | **4. 18252C>T (ORF1ab: N5995N)** | | | | |
| Lineage | Outbreak | Sample | A | C | G | T | num.reads | A | C | G | T | num.reads |
| B.1.1.33 | Rivera I | M5 | 1 | 19988 | 0 | 11 | 20000 | 32 | 11 | 1 | 20172 | 20216 |
| B.1.1.33 | Rivera I | M8 | 10 | 55176 | 2 | 1 | 55189 | 13 | 7 | 0 | 8994 | 9014 |
| B.1.1.33 | Rivera I | M9 | 1 | 25561 | 1 | 1 | 25564 | 22 | 7 | 2 | 9245 | 9276 |
| B.1.1.33 | Rivera I | M10 | 2 | 18667 | 1 | 10 | 18680 | 32 | 26 | 2 | 16098 | 16158 |
| B.1.1.33 | Rivera I | M11 | 0 | 18597 | 2 | 1 | 18600 | 28 | 12 | 0 | 17031 | 17071 |
| B.1.1.33 | Rivera I | M12 | 2 | 19479 | 0 | 0 | 19481 | 20 | 13 | 0 | 14090 | 14123 |
| B.1.1.33 | Rivera I | M13 | 1 | 17285 | 0 | 0 | 17286 | 31 | 24 | 0 | 16236 | 16291 |
| B.1.1.33 | Rivera I | M14 | 1 | 14015 | 1 | 0 | 14017 | 23 | 8 | 1 | 14182 | 14214 |
| B.1.1.33 | Rivera I | M15 | 1 | 19521 | 1 | 2 | 19525 | 16 | 15 | 2 | 18139 | 18172 |
| B.1.1.33 | Rivera I | M40 | 3 | 981 | 0 | 6 | 990 | 6 | 12 | 3 | 971 | 992 |
| B.1.1.33 | Rivera I | M43 | 1 | 982 | 2 | 8 | 993 | 3 | 21 | 5 | 949 | 978 |
| B.1.1.33 | Rivera I | M72 | 5 | 981 | 1 | 6 | 993 | 2 | 28 | 1 | 950 | 981 |
| B.1.1.33 | Rivera I | M73 | 2 | 983 | 0 | 9 | 994 | 1 | 22 | 1 | 810 | 834 |
| B.1.1.33 | Rivera I | M79 | 2 | 989 | 0 | 5 | 996 | 1 | 7 | 0 | 416 | 424 |
| B.1.1.33 | Rivera II | M119 | 1 | 13 | 1 | 267 | 282 | 0 | 271 | 3 | 8 | 282 |
| B.1.1.33 | Rivera II | M154 | 4 | 48 | 4 | 872 | 928 | 0 | 962 | 3 | 29 | 994 |
| B.1.1.33 | Rivera II | M155 | 8 | 49 | 2 | 879 | 938 | 0 | 618 | 4 | 21 | 643 |
| B.1.1.33 | | RS-15273 | 0 | 1200 | 0 | 3 | 1203 | 6 | 6 | 1 | 3110 | 3123 |
| B.1.1.33 | | RS-15280 | 0 | 49 | 0 | 0 | **49** | 1 | 455 | 1 | 38 | 495 |
| B.1.1.33 | | RS-6177 | 1 | 360 | 0 | 2 | 363 | 10 | 6371 | 6 | 10 | 6397 |
| B.1.1.33 | | RS-6188 | 0 | 46 | 0 | 1 | **47** | 2 | 2 | 0 | 546 | 550 |
| B.1.1.33 | | RS-6203 | 0 | 76 | 0 | 0 | **76** | 0 | 2 | 0 | 400 | 402 |
| B.1.1.33 | | RS-6208 | 0 | 55 | 0 | 0 | **55** | 0 | 128 | 1 | 1246 | 1375 |
| B.1.1.33 | | RS-6222 | 0 | 101 | 0 | 0 | 101 | 1 | 8 | 3 | 5403 | 5415 |
| B.1.1.33 | | RS-6231 | 0 | 43 | 0 | 0 | **43** | 8 | 8 | 10 | 7782 | 7808 |
| | | | **4. 18252C>T (ORF1ab: N5995N)** | | | | | **5. 23311G>T (Spike protein S1: E571D)** | | | | |
| Lineage | Outbreak | Sample | A | C | G | T | num.reads | A | C | G | T | num.reads |
| B.1.1.33 | Rivera I | M5 | 32 | 11 | 1 | 20172 | 20216 | 0 | 12 | 28247 | 15 | 28274 |
| B.1.1.33 | Rivera I | M8 | 13 | 7 | 0 | 8994 | 9014 | 1 | 0 | 56612 | 1 | 56614 |
| B.1.1.33 | Rivera I | M9 | 22 | 7 | 2 | 9245 | 9276 | 2 | 0 | 42747 | 3 | 42752 |
| B.1.1.33 | Rivera I | M10 | 32 | 26 | 2 | 16098 | 16158 | 1 | 0 | 22062 | 0 | 22063 |
| B.1.1.33 | Rivera I | M11 | 28 | 12 | 0 | 17031 | 17071 | 0 | 4 | 28490 | 0 | 28494 |
| B.1.1.33 | Rivera I | M12 | 20 | 13 | 0 | 14090 | 14123 | 0 | 0 | 22665 | 0 | 22665 |
| B.1.1.33 | Rivera I | M13 | 31 | 24 | 0 | 16236 | 16291 | 0 | 0 | 20148 | 3 | 20151 |
| B.1.1.33 | Rivera I | M14 | 23 | 8 | 1 | 14182 | 14214 | 0 | 0 | 15263 | 1 | 15264 |
| B.1.1.33 | Rivera I | M15 | 16 | 15 | 2 | 18139 | 18172 | 1 | 0 | 24879 | 1 | 24881 |
| B.1.1.33 | Rivera I | M40 | 6 | 12 | 3 | 971 | 992 | 13 | 1 | 948 | 1 | 963 |
| B.1.1.33 | Rivera I | M43 | 3 | 21 | 5 | 949 | 978 | 5 | 1 | 977 | 1 | 984 |
| B.1.1.33 | Rivera I | M72 | 2 | 28 | 1 | 950 | 981 | 3 | 4 | 983 | 1 | 991 |
| B.1.1.33 | Rivera I | M73 | 1 | 22 | 1 | 810 | 834 | 3 | 0 | 993 | 0 | 996 |
| B.1.1.33 | Rivera I | M79 | 1 | 7 | 0 | 416 | 424 | 2 | 0 | 988 | 0 | 990 |
| B.1.1.33 | Rivera II | M119 | 0 | 271 | 3 | 8 | 282 | 0 | 0 | 5 | 114 | 119 |
| B.1.1.33 | Rivera II | M154 | 0 | 962 | 3 | 29 | 994 | 0 | 0 | 2 | 83 | **85** |

| | | | | | | | | | | | | |
|---|---|---|---|---|---|---|---|---|---|---|---|---|
| B.1.1.33 | Rivera II | M155 | 0 | 618 | 4 | 21 | 643 | 4 | 1 | 17 | 972 | 994 |
| B.1.1.33 | | RS-15273 | 6 | 6 | 1 | 3110 | 3123 | 1 | 1 | 2631 | 5 | 2638 |
| B.1.1.33 | | RS-15280 | 1 | 455 | 1 | 38 | 495 | 5 | 1 | 488 | 1 | 495 |
| B.1.1.33 | | RS-6177 | 10 | 6371 | 6 | 10 | 6397 | 11 | 8 | 5405 | 14 | 5438 |
| B.1.1.33 | | RS-6188 | 2 | 2 | 0 | 546 | 550 | 0 | 3 | 1994 | 4 | 2001 |
| B.1.1.33 | | RS-6203 | 0 | 2 | 0 | 400 | 402 | 2 | 6 | 2649 | 6 | 2663 |
| B.1.1.33 | | RS-6208 | 0 | 128 | 1 | 1246 | 1375 | 1 | 1 | 807 | 0 | 809 |
| B.1.1.33 | | RS-6222 | 1 | 8 | 3 | 5403 | 5415 | 2 | 0 | 2277 | 3 | 2282 |
| B.1.1.33 | | RS-6231 | 8 | 8 | 10 | 7782 | 7808 | 8 | 3 | 2684 | 5 | 2700 |
| | | | **6. 24872G>T (Spike protein S2: V419L)** | | | | | **7. 28093C>T (ORF8: S52F)** | | | | |
| Lineage | Outbreak | Sample | A | C | G | T | num.reads | A | C | G | T | num.reads |
| B.1.1.33 | Rivera I | M5 | 0 | 1 | 3 | 19955 | 19959 | 0 | 14 | 0 | 6149 | 6163 |
| B.1.1.33 | Rivera I | M8 | 0 | 1 | 0 | 24167 | 24168 | 1 | 1 | 0 | 671 | 673 |
| B.1.1.33 | Rivera I | M9 | 0 | 0 | 3 | 38422 | 38425 | 0 | 3 | 0 | 1150 | 1153 |
| B.1.1.33 | Rivera I | M10 | 0 | 1 | 60 | 20587 | 20648 | 0 | 11 | 0 | 3753 | 3764 |
| B.1.1.33 | Rivera I | M11 | 0 | 1 | 0 | 17546 | 17547 | 0 | 6 | 0 | 3048 | 3054 |
| B.1.1.33 | Rivera I | M12 | 0 | 1 | 69 | 18470 | 18540 | 0 | 11 | 0 | 2389 | 2389 |
| B.1.1.33 | Rivera I | M13 | 2 | 16 | 4 | 18801 | 18823 | 0 | 6 | 0 | 3635 | 3641 |
| B.1.1.33 | Rivera I | M14 | 1 | 0 | 0 | 11543 | 11544 | 0 | 3 | 0 | 3477 | 3480 |
| B.1.1.33 | Rivera I | M15 | 0 | 0 | 3 | 20400 | 20403 | 1 | 11 | 0 | 2969 | 2981 |
| B.1.1.33 | Rivera I | M40 | 16 | 5 | 15 | 741 | 777 | 5 | 29 | 1 | 1779 | 1814 |
| B.1.1.33 | Rivera I | M43 | 13 | 11 | 10 | 767 | 801 | 0 | 4 | 0 | 163 | 167 |
| B.1.1.33 | Rivera I | M72 | 15 | 17 | 12 | 763 | 807 | 1 | 6 | 1 | 239 | 247 |
| B.1.1.33 | Rivera I | M73 | 10 | 11 | 15 | 782 | 818 | 0 | 1 | 0 | 95 | **96** |
| B.1.1.33 | Rivera I | M79 | 18 | 13 | 13 | 766 | 810 | 0 | 0 | 0 | 100 | 100 |
| B.1.1.33 | Rivera II | M119 | 3 | 1 | 205 | 0 | 209 | 0 | 9 | 1 | 381 | 391 |
| B.1.1.33 | Rivera II | M154 | 22 | 4 | 959 | 2 | 987 | 0 | 3 | 3 | 323 | 329 |
| B.1.1.33 | Rivera II | M155 | 16 | 1 | 959 | 5 | 981 | 0 | 1 | 1 | 176 | 178 |
| B.1.1.33 | | RS-15273 | 1 | 0 | 1619 | 0 | 1620 | 3 | 0 | 0 | 2590 | 2593 |
| B.1.1.33 | | RS-15280 | 10 | 2 | 226 | 0 | 238 | 1 | 19 | 1 | 492 | 513 |
| B.1.1.33 | | RS-6177 | 0 | 1 | 2161 | 3 | 2165 | 4 | 3 | 6 | 4005 | 4018 |
| B.1.1.33 | | RS-6188 | 0 | 0 | 0 | 249 | 249 | 1 | 9 | 1 | 3258 | 3269 |
| B.1.1.33 | | RS-6203 | 0 | 0 | 309 | 2 | 311 | 0 | 3 | 3 | 3495 | 3501 |
| B.1.1.33 | | RS-6208 | 1 | 0 | 324 | 1 | 326 | 0 | 190 | 4 | 1046 | 1240 |
| B.1.1.33 | | RS-6222 | 1 | 0 | 2656 | 0 | 2657 | 1 | 5 | 2 | 2473 | 2481 |
| B.1.1.33 | | RS-6231 | 0 | 0 | 1567 | 1 | 1568 | 2 | 7 | 3 | 1871 | 1883 |
| **for comparison: CLADE BR-UY-I33** | | | | | | | | | | | | |
| | | | **1. 189G>T (5'-UTR)** | | | | | **2. 2276A>G (ORF1ab: I671V)** | | | | |
| Lineage | Outbreak | Sample | A | C | G | T | num.reads | A | C | G | T | num.reads |
| B.1.1.33 | Treinta y Tres | M49 | 0 | 1 | 131 | 0 | 132 | 985 | 3 | 5 | 0 | 993 |
| B.1.1.33 | Treinta y Tres | M52 | 12 | 3 | 970 | 1 | 986 | 63 | 0 | 3 | 1 | 67 |

| | | | | | | | | | | | | |
|---|---|---|---|---|---|---|---|---|---|---|---|---|
| B.1.1.33 | Treinta y Tres | M53 | 6 | 1 | 662 | 2 | 671 | 4 | 0 | 0 | 0 | 4 |
| B.1.1.33 | Treinta y Tres | M54 | 1 | 0 | 269 | 0 | 270 | 52 | 0 | 1 | 0 | 53 |
| B.1.1.33 | Treinta y Tres | M57 | 3 | 0 | 313 | 0 | 316 | 976 | 1 | 7 | 1 | 985 |
| B.1.1.33 | Treinta y Tres | M59 | 6 | 2 | 986 | 1 | 995 | 6 | 0 | 0 | 0 | 6 |
| B.1.1.33 | Treinta y Tres | M60 | 9 | 2 | 765 | 2 | 778 | 13 | 0 | 1 | 0 | 14 |
| B.1.1.33 | Treinta y Tres | M62 | 10 | 0 | 584 | 1 | 595 | 18 | 0 | 0 | 0 | 18 |
| B.1.1.33 | Treinta y Tres | M63 | 12 | 1 | 972 | 1 | 986 | 16 | 0 | 0 | 0 | 16 |
| B.1.1.33 | Treinta y Tres | M64 | 6 | 1 | 411 | 0 | 418 | 985 | 1 | 6 | 2 | 994 |
| B.1.1.33 | Treinta y Tres | M65 | 0 | 0 | 2 | 0 | 2 | 379 | 0 | 6 | 0 | 385 |
| B.1.1.33 | Treinta y Tres | M66 | 0 | 0 | 0 | 0 | 0 | 577 | 1 | 10 | 2 | 590 |
| B.1.1.33 | Treinta y Tres | M67 | 0 | 0 | 0 | 0 | 0 | 449 | 0 | 7 | 0 | 456 |
| B.1.1.33 | Treinta y Tres | M68 | 0 | 0 | 7 | 0 | 7 | 505 | 1 | 9 | 0 | 515 |
| B.1.1.33 | Treinta y Tres | M69 | 0 | 0 | 1 | 1 | 2 | 703 | 2 | 11 | 1 | 717 |
| B.1.1.33 | Treinta y Tres | M70 | 0 | 0 | 4 | 0 | 4 | 976 | 0 | 12 | 1 | 989 |
| B.1.1.33 | Treinta y Tres | M71 | 0 | 0 | 2 | 0 | 2 | 483 | 0 | 7 | 0 | 490 |
| B.1.1.33 | Treinta y Tres | M85 | 12 | 0 | 978 | 1 | 991 | 990 | 0 | 3 | 0 | 993 |
| B.1.1.33 | Treinta y Tres | M87 | 15 | 0 | 966 | 4 | 985 | 973 | 2 | 6 | 1 | 982 |
| B.1.1.33 | Treinta y Tres | M91 | 8 | 0 | 916 | 1 | 925 | 978 | 1 | 10 | 0 | 989 |
| B.1.1.33 | Treinta y Tres | M93 | 4 | 2 | 982 | 2 | 990 | 981 | 3 | 7 | 2 | 993 |
| B.1.1.33 | Treinta y Tres | M94 | 8 | 1 | 979 | 0 | 988 | 410 | 0 | 3 | 0 | 413 |
| B.1.1.33 | Treinta y Tres | M95 | 11 | 1 | 971 | 2 | 985 | 253 | 0 | 2 | 0 | 255 |
| B.1.1.33 | Treinta y Tres | M97 | 3 | 0 | 234 | 0 | 237 | 984 | 0 | 8 | 0 | 992 |
| B.1.1.33 | Treinta y Tres | M98 | 5 | 2 | 974 | 1 | 982 | 210 | 0 | 0 | 0 | 210 |
| B.1.1.33 | Treinta y Tres | M130 | 10 | 1 | 978 | 3 | 992 | 124 | 0 | 0 | 0 | 124 |
| B.1.1.33 | Treinta y Tres | M132 | 8 | 2 | 974 | 0 | 984 | 417 | 0 | 4 | 1 | 422 |
| B.1.1.33 | Treinta y Tres | M133 | 8 | 3 | 816 | 1 | 828 | 772 | 1 | 10 | 1 | 784 |
| B.1.1.33 | Treinta y Tres | M135 | 3 | 2 | 569 | 2 | 576 | 695 | 0 | 6 | 2 | 703 |
| B.1.1.33 | Treinta y Tres | M136 | 1 | 0 | 103 | 0 | 104 | 974 | 2 | 7 | 0 | 983 |
| B.1.1.33 | | RJ-2007 | 2 | 2 | 486 | 3 | 493 | 486 | 1 | 7 | 0 | 494 |
| B.1.1.33 | | RJ-2091 | 0 | 1 | 500 | 0 | 501 | 605 | 0 | 0 | 0 | 605 |
| B.1.1.33 | | RJ-2342 | 1 | 0 | 389 | 1 | 391 | 645 | 0 | 0 | 1 | 646 |
| | | | **3. 12025C>T (ORF1ab: S3920S)** | | | | | **4. 18252C>T (ORF1ab: N5995N)** | | | | |
| Lineage | Outbreak | Sample | A | C | G | T | num.reads | A | C | G | T | num.reads |
| B.1.1.33 | Treinta y Tres | M49 | 4 | 767 | 0 | 4 | 775 | 1 | 821 | 3 | 25 | 850 |
| B.1.1.33 | Treinta y Tres | M52 | 6 | 974 | 1 | 5 | 986 | 2 | 566 | 5 | 13 | 586 |
| B.1.1.33 | Treinta y Tres | M53 | 0 | 194 | 0 | 1 | 195 | 0 | 24 | 0 | 1 | 25 |
| B.1.1.33 | Treinta y Tres | M54 | 2 | 784 | 1 | 3 | 790 | 1 | 343 | 0 | 6 | 350 |

| | | | | | | | | | | | | |
|---|---|---|---|---|---|---|---|---|---|---|---|---|
| B.1.1.33 | Treinta y Tres | M57 | 1 | 780 | 0 | 3 | 784 | 0 | 702 | 4 | 22 | 728 |
| B.1.1.33 | Treinta y Tres | M59 | 0 | 172 | 0 | 1 | 173 | 0 | 32 | 0 | 2 | 34 |
| B.1.1.33 | Treinta y Tres | M60 | 0 | 524 | 1 | 4 | 529 | 0 | 79 | 0 | 1 | 80 |
| B.1.1.33 | Treinta y Tres | M62 | 6 | 838 | 0 | 1 | 845 | 0 | 159 | 0 | 2 | 161 |
| B.1.1.33 | Treinta y Tres | M63 | 3 | 518 | 1 | 7 | 529 | 0 | 50 | 0 | 0 | 50 |
| B.1.1.33 | Treinta y Tres | M64 | 3 | 906 | 0 | 4 | 913 | 0 | 967 | 3 | 20 | 990 |
| B.1.1.33 | Treinta y Tres | M65 | 2 | 683 | 2 | 3 | 690 | 0 | 827 | 5 | 27 | 859 |
| B.1.1.33 | Treinta y Tres | M66 | 2 | 654 | 0 | 6 | 662 | 0 | 877 | 6 | 24 | 907 |
| B.1.1.33 | Treinta y Tres | M67 | 1 | 790 | 0 | 3 | 794 | 0 | 873 | 10 | 22 | 905 |
| B.1.1.33 | Treinta y Tres | M68 | 2 | 375 | 0 | 0 | 377 | 1 | 751 | 2 | 23 | 777 |
| B.1.1.33 | Treinta y Tres | M69 | 2 | 648 | 0 | 3 | 653 | 1 | 955 | 2 | 29 | 987 |
| B.1.1.33 | Treinta y Tres | M70 | 2 | 982 | 2 | 4 | 990 | 0 | 943 | 5 | 39 | 987 |
| B.1.1.33 | Treinta y Tres | M71 | 3 | 762 | 2 | 4 | 771 | 0 | 956 | 6 | 31 | 993 |
| B.1.1.33 | Treinta y Tres | M85 | 1 | 981 | 0 | 5 | 987 | 0 | 967 | 3 | 25 | 995 |
| B.1.1.33 | Treinta y Tres | M87 | 5 | 972 | 2 | 6 | 985 | 1 | 952 | 4 | 25 | 982 |
| B.1.1.33 | Treinta y Tres | M91 | 1 | 983 | 0 | 6 | 990 | 2 | 957 | 6 | 28 | 993 |
| B.1.1.33 | Treinta y Tres | M93 | 3 | 980 | 1 | 6 | 990 | 1 | 963 | 3 | 21 | 988 |
| B.1.1.33 | Treinta y Tres | M94 | 1 | 981 | 0 | 8 | 990 | 1 | 950 | 3 | 40 | 994 |
| B.1.1.33 | Treinta y Tres | M95 | 1 | 983 | 0 | 5 | 989 | 0 | 966 | 2 | 20 | 988 |
| B.1.1.33 | Treinta y Tres | M97 | 4 | 980 | 1 | 3 | 988 | 0 | 960 | 3 | 28 | 991 |
| B.1.1.33 | Treinta y Tres | M98 | 2 | 990 | 0 | 6 | 998 | 0 | 972 | 6 | 15 | 993 |
| B.1.1.33 | Treinta y Tres | M130 | 2 | 985 | 1 | 7 | 995 | 0 | 951 | 4 | 20 | 975 |
| B.1.1.33 | Treinta y Tres | M132 | 2 | 982 | 0 | 5 | 989 | 0 | 958 | 7 | 29 | 994 |
| B.1.1.33 | Treinta y Tres | M133 | 0 | 970 | 0 | 9 | 979 | 2 | 947 | 7 | 33 | 989 |
| B.1.1.33 | Treinta y Tres | M135 | 3 | 739 | 2 | 10 | 754 | 0 | 591 | 3 | 17 | 611 |
| B.1.1.33 | Treinta y Tres | M136 | 0 | 289 | 0 | 4 | 293 | 0 | 193 | 3 | 4 | 200 |
| B.1.1.33 | | RJ-2007 | 0 | 185 | 0 | 0 | 185 | 0 | 461 | 4 | 28 | 493 |
| B.1.1.33 | | RJ-2091 | 0 | 396 | 0 | 2 | 398 | 1 | 1507 | 0 | 1 | 1509 |
| B.1.1.33 | | RJ-2342 | 1 | 215 | 0 | 2 | 218 | 8 | 4432 | 1 | 6 | 4447 |
| | | | **5. 23311G>T (Spike protein S1: E571D)** | | | | | **6. 24872G>T (Spike protein S2: V419L)** | | | | |
| Lineage | Outbreak | Sample | A | C | G | T | num.reads | A | C | G | T | num.reads |
| B.1.1.33 | Treinta y Tres | M49 | 2 | 0 | 893 | 1 | 896 | 3 | 0 | 303 | 5 | 311 |
| B.1.1.33 | Treinta y Tres | M52 | 4 | 1 | 971 | 2 | 978 | 21 | 3 | 961 | 2 | 987 |
| B.1.1.33 | Treinta y Tres | M53 | 7 | 1 | 982 | 0 | 990 | 13 | 0 | 333 | 1 | 347 |
| B.1.1.33 | Treinta y Tres | M54 | 3 | 0 | 987 | 1 | 991 | 22 | 8 | 951 | 2 | 983 |
| B.1.1.33 | Treinta y Tres | M57 | 6 | 2 | 980 | 3 | 991 | 10 | 0 | 544 | 1 | 555 |
| B.1.1.33 | Treinta y Tres | M59 | 4 | 0 | 986 | 0 | 990 | 18 | 0 | 644 | 3 | 665 |

| | | | | | | | | | | | | |
|---|---|---|---|---|---|---|---|---|---|---|---|---|
| B.1.1.33 | Treinta y Tres | M60 | 5 | 1 | 981 | 1 | 988 | 34 | 5 | 779 | 7 | 825 |
| B.1.1.33 | Treinta y Tres | M62 | 3 | 0 | 987 | 3 | 993 | 25 | 3 | 927 | 3 | 958 |
| B.1.1.33 | Treinta y Tres | M63 | 9 | 1 | 982 | 3 | 995 | 25 | 4 | 955 | 0 | 984 |
| B.1.1.33 | Treinta y Tres | M64 | 7 | 3 | 980 | 0 | 990 | 7 | 1 | 559 | 1 | 568 |
| B.1.1.33 | Treinta y Tres | M65 | 7 | 1 | 986 | 0 | 994 | 19 | 3 | 622 | 2 | 646 |
| B.1.1.33 | Treinta y Tres | M66 | 3 | 4 | 982 | 2 | 991 | 19 | 1 | 645 | 1 | 666 |
| B.1.1.33 | Treinta y Tres | M67 | 2 | 1 | 743 | 0 | 746 | 13 | 2 | 408 | 0 | 423 |
| B.1.1.33 | Treinta y Tres | M68 | 12 | 1 | 950 | 3 | 966 | 13 | 4 | 480 | 4 | 501 |
| B.1.1.33 | Treinta y Tres | M69 | 5 | 4 | 977 | 1 | 987 | 15 | 3 | 665 | 5 | 688 |
| B.1.1.33 | Treinta y Tres | M70 | 4 | 2 | 974 | 1 | 981 | 21 | 3 | 964 | 0 | 988 |
| B.1.1.33 | Treinta y Tres | M71 | 6 | 2 | 982 | 3 | 993 | 26 | 7 | 591 | 4 | 628 |
| B.1.1.33 | Treinta y Tres | M85 | 5 | 1 | 987 | 0 | 993 | 18 | 4 | 960 | 3 | 985 |
| B.1.1.33 | Treinta y Tres | M87 | 5 | 3 | 979 | 0 | 987 | 14 | 4 | 958 | 7 | 983 |
| B.1.1.33 | Treinta y Tres | M91 | 2 | 3 | 849 | 1 | 855 | 19 | 0 | 962 | 4 | 985 |
| B.1.1.33 | Treinta y Tres | M93 | 3 | 2 | 984 | 0 | 989 | 17 | 1 | 965 | 5 | 988 |
| B.1.1.33 | Treinta y Tres | M94 | 5 | 1 | 977 | 0 | 983 | 13 | 6 | 960 | 4 | 983 |
| B.1.1.33 | Treinta y Tres | M95 | 2 | 1 | 987 | 4 | 994 | 21 | 8 | 954 | 3 | 986 |
| B.1.1.33 | Treinta y Tres | M97 | 4 | 3 | 807 | 1 | 815 | 9 | 1 | 339 | 0 | 349 |
| B.1.1.33 | Treinta y Tres | M98 | 2 | 0 | 993 | 1 | 996 | 15 | 3 | 963 | 4 | 985 |
| B.1.1.33 | Treinta y Tres | M130 | 3 | 1 | 984 | 0 | 988 | 19 | 4 | 966 | 1 | 990 |
| B.1.1.33 | Treinta y Tres | M132 | 8 | 0 | 984 | 0 | 992 | 22 | 2 | 962 | 3 | 989 |
| B.1.1.33 | Treinta y Tres | M133 | 0 | 0 | 148 | 0 | 148 | 12 | 5 | 906 | 6 | 929 |
| B.1.1.33 | Treinta y Tres | M135 | 1 | 0 | 155 | 0 | 156 | 9 | 0 | 409 | 2 | 420 |
| B.1.1.33 | Treinta y Tres | M136 | 1 | 0 | 170 | 0 | 171 | 1 | 1 | 261 | 1 | 264 |
| B.1.1.33 | | RJ-2007 | 1 | 1 | 489 | 1 | 492 | 7 | 3 | 481 | 1 | 492 |
| B.1.1.33 | | RJ-2091 | 1 | 0 | 1180 | 2 | 1183 | 0 | 0 | 612 | 0 | 612 |
| B.1.1.33 | | RJ-2342 | 4 | 1 | 3112 | 7 | 3124 | 0 | 0 | 1572 | 1 | 1573 |
| | | | **7. 28093C>T (ORF8: S52F)** | | | | | | | | | |
| Lineage | Outbreak | Sample | A | C | G | T | num.reads | | | | | |
| B.1.1.33 | Treinta y Tres | M49 | 5 | 1951 | 2 | 12 | 1970 | | | | | |
| B.1.1.33 | Treinta y Tres | M52 | 0 | 115 | 0 | 0 | 115 | | | | | |
| B.1.1.33 | Treinta y Tres | M53 | 0 | 11 | 0 | 0 | 11 | | | | | |
| B.1.1.33 | Treinta y Tres | M54 | 0 | 129 | 0 | 1 | 130 | | | | | |
| B.1.1.33 | Treinta y Tres | M57 | 3 | 1888 | 2 | 19 | 1912 | | | | | |
| B.1.1.33 | Treinta y Tres | M59 | 0 | 11 | 0 | 0 | 11 | | | | | |
| B.1.1.33 | Treinta y Tres | M60 | 0 | 23 | 0 | 0 | 23 | | | | | |
| B.1.1.33 | Treinta y Tres | M62 | 0 | 18 | 0 | 0 | 18 | | | | | |

| | | | | | | | |
|---|---|---|---|---|---|---|---|
| B.1.1.33 | Treinta y Tres | M63 | 0 | 30 | 0 | 0 | 30 |
| B.1.1.33 | Treinta y Tres | M64 | 1 | 1937 | 1 | 25 | 1964 |
| B.1.1.33 | Treinta y Tres | M65 | 0 | 494 | 1 | 13 | 508 |
| B.1.1.33 | Treinta y Tres | M66 | 1 | 511 | 0 | 5 | 517 |
| B.1.1.33 | Treinta y Tres | M67 | 0 | 655 | 1 | 6 | 662 |
| B.1.1.33 | Treinta y Tres | M68 | 1 | 396 | 0 | 3 | 400 |
| B.1.1.33 | Treinta y Tres | M69 | 0 | 609 | 1 | 7 | 617 |
| B.1.1.33 | Treinta y Tres | M70 | 2 | 972 | 0 | 9 | 983 |
| B.1.1.33 | Treinta y Tres | M71 | 0 | 494 | 1 | 10 | 505 |
| B.1.1.33 | Treinta y Tres | M85 | 2 | 980 | 0 | 2 | 984 |
| B.1.1.33 | Treinta y Tres | M87 | 1 | 975 | 0 | 10 | 986 |
| B.1.1.33 | Treinta y Tres | M91 | 2 | 1308 | 0 | 14 | 1324 |
| B.1.1.33 | Treinta y Tres | M93 | 0 | 980 | 0 | 13 | 993 |
| B.1.1.33 | Treinta y Tres | M94 | 1 | 984 | 0 | 5 | 990 |
| B.1.1.33 | Treinta y Tres | M95 | 3 | 980 | 0 | 6 | 989 |
| B.1.1.33 | Treinta y Tres | M97 | 2 | 1958 | 0 | 25 | 1985 |
| B.1.1.33 | Treinta y Tres | M98 | 3 | 970 | 0 | 11 | 984 |
| B.1.1.33 | Treinta y Tres | M130 | 0 | 543 | 1 | 8 | 552 |
| B.1.1.33 | Treinta y Tres | M132 | 3 | 859 | 3 | 7 | 872 |
| B.1.1.33 | Treinta y Tres | M133 | 0 | 482 | 1 | 10 | 493 |
| B.1.1.33 | Treinta y Tres | M135 | 3 | 539 | 2 | 3 | 547 |
| B.1.1.33 | Treinta y Tres | M136 | 1 | 980 | 0 | 10 | 991 |
| B.1.1.33 | | RJ-2007 | 4 | 973 | 5 | 10 | 992 |
| B.1.1.33 | | RJ-2091 | 5 | 1371 | 0 | 3 | 1379 |
| B.1.1.33 | | RJ-2342 | 5 | 1776 | 0 | 1 | 1782 |

**Table S5.** Within-host diversity BR-UY-I33. The table shows the observed allele count for each synapomorphic site identified in clade BR-UY-I33

| **CLADE BR-UY-I33** | | | | | | | | | | | | |
|---|---|---|---|---|---|---|---|---|---|---|---|---|
| | | | **1. 5089A>G (ORF1ab: K1608K)** | | | | | **2. 8293C>T (ORF1ab: T2676T)** | | | | |
| Lineage | Outbreak | Sample | A | C | G | T | num.reads | A | C | G | T | num.reads |
| B.1.1.33 | Treinta y Tres | M49 | 82 | 5 | 469 | 4 | 560 | 2 | 12 | 0 | 289 | 303 |
| B.1.1.33 | Treinta y Tres | M52 | 3 | 0 | 10 | 0 | 13 | 0 | 4 | 0 | 30 | 34 |
| B.1.1.33 | Treinta y Tres | M53 | 1 | 0 | 3 | 0 | 4 | 0 | 0 | 0 | 46 | 46 |
| B.1.1.33 | Treinta y Tres | M54 | 0 | 0 | 6 | 0 | 6 | 0 | 3 | 0 | 34 | 37 |
| B.1.1.33 | Treinta y Tres | M57 | 95 | 7 | 397 | 3 | 502 | 2 | 28 | 0 | 534 | 564 |
| B.1.1.33 | Treinta y Tres | M59 | 2 | 0 | 4 | 0 | 6 | 0 | 4 | 0 | 56 | 60 |
| B.1.1.33 | Treinta y Tres | M60 | 0 | 0 | 7 | 0 | 7 | 0 | 2 | 0 | 40 | 42 |

| | | | | | | | | | | | | |
|---|---|---|---|---|---|---|---|---|---|---|---|---|
| B.1.1.33 | Treinta y Tres | M62 | 2 | 0 | 5 | 0 | 7 | 0 | 2 | 0 | 38 | 40 |
| B.1.1.33 | Treinta y Tres | M63 | 1 | 0 | 11 | 1 | 13 | 0 | 2 | 0 | 73 | 75 |
| B.1.1.33 | Treinta y Tres | M64 | 157 | 15 | 817 | 5 | 994 | 6 | 47 | 1 | 797 | 851 |
| B.1.1.33 | Treinta y Tres | M65 | 90 | 5 | 395 | 3 | 493 | 16 | 82 | 1 | 1361 | 1460 |
| B.1.1.33 | Treinta y Tres | M66 | 32 | 4 | 84 | 2 | 122 | 7 | 69 | 4 | 1166 | 1246 |
| B.1.1.33 | Treinta y Tres | M67 | 26 | 1 | 105 | 0 | 132 | 4 | 90 | 0 | 1407 | 1501 |
| B.1.1.33 | Treinta y Tres | M68 | 59 | 7 | 266 | 5 | 337 | 6 | 108 | 9 | 1454 | 1577 |
| B.1.1.33 | Treinta y Tres | M69 | 126 | 13 | 457 | 10 | 606 | 8 | 83 | 2 | 1697 | 1790 |
| B.1.1.33 | Treinta y Tres | M70 | 74 | 3 | 329 | 2 | 408 | 13 | 92 | 0 | 1788 | 1893 |
| B.1.1.33 | Treinta y Tres | M71 | 44 | 8 | 148 | 1 | 201 | 11 | 93 | 6 | 1529 | 1639 |
| B.1.1.33 | Treinta y Tres | M85 | 145 | 10 | 828 | 6 | 989 | 8 | 49 | 0 | 929 | 986 |
| B.1.1.33 | Treinta y Tres | M87 | 160 | 16 | 792 | 7 | 975 | 20 | 101 | 6 | 1828 | 1955 |
| B.1.1.33 | Treinta y Tres | M91 | 142 | 7 | 826 | 8 | 983 | 6 | 46 | 1 | 928 | 981 |
| B.1.1.33 | Treinta y Tres | M93 | 166 | 20 | 805 | 4 | 995 | 13 | 9 | 2 | 1856 | 1880 |
| B.1.1.33 | Treinta y Tres | M94 | 155 | 15 | 819 | 6 | 995 | 9 | 87 | 2 | 1738 | 1836 |
| B.1.1.33 | Treinta y Tres | M95 | 148 | 16 | 829 | 2 | 995 | 2 | 69 | 0 | 1360 | 1431 |
| B.1.1.33 | Treinta y Tres | M97 | 156 | 10 | 815 | 7 | 988 | 10 | 40 | 2 | 826 | 878 |
| B.1.1.33 | Treinta y Tres | M98 | 170 | 10 | 784 | 6 | 970 | 9 | 72 | 2 | 1292 | 1375 |
| B.1.1.33 | Treinta y Tres | M130 | 2 | 1 | 20 | 0 | 23 | 0 | 8 | 0 | 202 | 210 |
| B.1.1.33 | Treinta y Tres | M132 | 7 | 0 | 32 | 1 | 40 | 3 | 57 | 0 | 794 | 854 |
| B.1.1.33 | Treinta y Tres | M133 | 146 | 14 | 693 | 6 | 859 | 6 | 59 | 1 | 902 | 968 |
| B.1.1.33 | Treinta y Tres | M135 | 57 | 10 | 320 | 5 | 392 | 1 | 28 | 0 | 506 | 535 |
| B.1.1.33 | Treinta y Tres | M136 | 21 | 6 | 115 | 0 | 142 | 2 | 7 | 0 | 125 | 134 |
| B.1.1.33 | | RJ-2007 | 489 | 4 | 1 | 2 | 496 | 3 | 490 | 0 | 5 | 498 |
| B.1.1.33 | | RJ-2091 | 402 | 3 | 2 | 3 | 410 | 0 | 976 | 1 | 2 | 979 |
| B.1.1.33 | | RJ-2342 | 639 | 0 | 1 | 2 | 642 | 5 | 4335 | 2 | 3 | 4345 |
| | | | **3. 15108C>T (ORF1ab: T4947T)** | | | | | **4. 22720A>G (Spike protein S1: K374K)** | | | | |
| Lineage | Outbreak | Sample | A | C | G | T | num.reads | A | C | G | T | num.reads |
| B.1.1.33 | Treinta y Tres | M49 | 6 | 27 | 2 | 908 | 943 | 37 | 1 | 749 | 7 | 794 |
| B.1.1.33 | Treinta y Tres | M52 | 4 | 79 | 5 | 845 | 933 | 50 | 2 | 934 | 7 | 993 |
| B.1.1.33 | Treinta y Tres | M53 | 1 | 1 | 0 | 40 | 42 | 14 | 0 | 327 | 2 | 343 |
| B.1.1.33 | Treinta y Tres | M54 | 6 | 25 | 0 | 452 | 483 | 34 | 0 | 711 | 4 | 749 |
| B.1.1.33 | Treinta y Tres | M57 | 5 | 54 | 0 | 927 | 986 | 45 | 2 | 734 | 5 | 786 |
| B.1.1.33 | Treinta y Tres | M59 | 1 | 5 | 0 | 63 | 69 | 39 | 1 | 701 | 6 | 747 |
| B.1.1.33 | Treinta y Tres | M60 | 0 | 4 | 0 | 102 | 106 | 31 | 2 | 638 | 3 | 674 |
| B.1.1.33 | Treinta y Tres | M62 | 0 | 12 | 0 | 113 | 125 | 39 | 1 | 833 | 3 | 876 |
| B.1.1.33 | Treinta y Tres | M63 | 2 | 4 | 0 | 138 | 144 | 66 | 1 | 921 | 2 | 990 |
| B.1.1.33 | Treinta y Tres | M64 | 4 | 59 | 4 | 919 | 986 | 49 | 3 | 901 | 3 | 956 |
| B.1.1.33 | Treinta y Tres | M65 | 2 | 27 | 1 | 273 | 303 | 54 | 4 | 908 | 4 | 970 |
| B.1.1.33 | Treinta y Tres | M66 | 1 | 16 | 2 | 281 | 300 | 52 | 0 | 737 | 5 | 794 |
| B.1.1.33 | Treinta y Tres | M67 | 1 | 7 | 1 | 98 | 107 | 55 | 3 | 921 | 7 | 986 |
| B.1.1.33 | Treinta y Tres | M68 | 6 | 38 | 4 | 515 | 563 | 58 | 4 | 888 | 7 | 957 |
| B.1.1.33 | Treinta y Tres | M69 | 6 | 60 | 8 | 816 | 890 | 60 | 4 | 924 | 2 | 990 |
| B.1.1.33 | Treinta y Tres | M70 | 2 | 466 | 0 | 13 | 481 | 55 | 2 | 925 | 11 | 993 |
| B.1.1.33 | Treinta y Tres | M71 | 5 | 25 | 1 | 360 | 391 | 60 | 5 | 921 | 3 | 989 |
| B.1.1.33 | Treinta y Tres | M85 | 1 | 71 | 1 | 913 | 986 | 51 | 1 | 931 | 4 | 987 |
| B.1.1.33 | Treinta y Tres | M87 | 3 | 55 | 2 | 925 | 985 | 65 | 0 | 912 | 6 | 983 |
| B.1.1.33 | Treinta y Tres | M91 | 4 | 43 | 3 | 931 | 981 | 54 | 0 | 922 | 4 | 980 |
| B.1.1.33 | Treinta y Tres | M93 | 2 | 66 | 0 | 911 | 979 | 65 | 2 | 917 | 5 | 989 |
| B.1.1.33 | Treinta y Tres | M94 | 6 | 64 | 7 | 909 | 986 | 43 | 1 | 941 | 6 | 991 |
| B.1.1.33 | Treinta y Tres | M95 | 3 | 41 | 1 | 932 | 977 | 54 | 2 | 932 | 2 | 990 |

| | | | | | | | | | | | | |
|---|---|---|---|---|---|---|---|---|---|---|---|---|
| B.1.1.33 | Treinta y Tres | M97 | 3 | 52 | 6 | 921 | 982 | 51 | 0 | 933 | 5 | 989 |
| B.1.1.33 | Treinta y Tres | M98 | 3 | 52 | 2 | 926 | 983 | 47 | 3 | 939 | 6 | 995 |
| B.1.1.33 | Treinta y Tres | M130 | 8 | 76 | 2 | 823 | 909 | 57 | 3 | 924 | 6 | 990 |
| B.1.1.33 | Treinta y Tres | M132 | 7 | 79 | 3 | 888 | 977 | 69 | 1 | 917 | 5 | 992 |
| B.1.1.33 | Treinta y Tres | M133 | 0 | 9 | 0 | 126 | 135 | 23 | 1 | 565 | 5 | 594 |
| B.1.1.33 | Treinta y Tres | M135 | 1 | 13 | 0 | 111 | 125 | 22 | 0 | 277 | 1 | 300 |
| B.1.1.33 | Treinta y Tres | M136 | 1 | 15 | 1 | 166 | 183 | 7 | 1 | 200 | 0 | 208 |
| B.1.1.33 | | RJ-2007 | 0 | 464 | 3 | 31 | 498 | 37 | 0 | 454 | 3 | 494 |
| B.1.1.33 | | RJ-2091 | 3 | 1409 | 6 | 4 | 1422 | 0 | 0 | 215 | 0 | 215 |
| B.1.1.33 | | RJ-2342 | 2 | 2390 | 2 | 3 | 2397 | 2 | 0 | 462 | 2 | 466 |
| | | | **5. 28775C>T (N: P168S)** | | | | | | | | | |
| Lineage | Outbreak | Sample | A | C | G | T | num.reads | | | | | |
| B.1.1.33 | Treinta y Tres | M49 | 5 | 53 | 3 | 924 | 985 | | | | | |
| B.1.1.33 | Treinta y Tres | M52 | 0 | 16 | 1 | 233 | 250 | | | | | |
| B.1.1.33 | Treinta y Tres | M53 | 3 | 14 | 1 | 200 | 218 | | | | | |
| B.1.1.33 | Treinta y Tres | M54 | 1 | 16 | 0 | 314 | 331 | | | | | |
| B.1.1.33 | Treinta y Tres | M57 | 8 | 49 | 1 | 926 | 984 | | | | | |
| B.1.1.33 | Treinta y Tres | M59 | 0 | 5 | 1 | 95 | 101 | | | | | |
| B.1.1.33 | Treinta y Tres | M60 | 2 | 7 | 0 | 191 | 200 | | | | | |
| B.1.1.33 | Treinta y Tres | M62 | 4 | 25 | 0 | 299 | 328 | | | | | |
| B.1.1.33 | Treinta y Tres | M63 | 4 | 24 | 2 | 352 | 382 | | | | | |
| B.1.1.33 | Treinta y Tres | M64 | 7 | 64 | 2 | 913 | 986 | | | | | |
| B.1.1.33 | Treinta y Tres | M65 | 2 | 21 | 3 | 290 | 316 | | | | | |
| B.1.1.33 | Treinta y Tres | M66 | 0 | 1 | 0 | 52 | 53 | | | | | |
| B.1.1.33 | Treinta y Tres | M67 | 1 | 3 | 0 | 24 | 28 | | | | | |
| B.1.1.33 | Treinta y Tres | M68 | 3 | 25 | 1 | 293 | 322 | | | | | |
| B.1.1.33 | Treinta y Tres | M69 | 5 | 39 | 2 | 453 | 499 | | | | | |
| B.1.1.33 | Treinta y Tres | M70 | 1 | 10 | 0 | 22 | 33 | | | | | |
| B.1.1.33 | Treinta y Tres | M71 | 1 | 12 | 0 | 161 | 174 | | | | | |
| B.1.1.33 | Treinta y Tres | M85 | 6 | 40 | 1 | 744 | 791 | | | | | |
| B.1.1.33 | Treinta y Tres | M87 | 5 | 70 | 4 | 904 | 983 | | | | | |
| B.1.1.33 | Treinta y Tres | M91 | 3 | 20 | 0 | 385 | 408 | | | | | |
| B.1.1.33 | Treinta y Tres | M93 | 7 | 41 | 0 | 934 | 982 | | | | | |
| B.1.1.33 | Treinta y Tres | M94 | 4 | 53 | 4 | 932 | 993 | | | | | |
| B.1.1.33 | Treinta y Tres | M95 | 2 | 22 | 0 | 312 | 336 | | | | | |
| B.1.1.33 | Treinta y Tres | M97 | 7 | 63 | 3 | 912 | 985 | | | | | |
| B.1.1.33 | Treinta y Tres | M98 | 6 | 33 | 1 | 550 | 590 | | | | | |
| B.1.1.33 | Treinta y Tres | M130 | 0 | 13 | 0 | 162 | 175 | | | | | |
| B.1.1.33 | Treinta y Tres | M132 | 0 | 13 | 1 | 171 | 185 | | | | | |
| B.1.1.33 | Treinta y Tres | M133 | 5 | 27 | 0 | 472 | 504 | | | | | |
| B.1.1.33 | Treinta y Tres | M135 | 4 | 36 | 0 | 638 | 678 | | | | | |
| B.1.1.33 | Treinta y Tres | M136 | 2 | 35 | 2 | 479 | 518 | | | | | |
| B.1.1.33 | | RJ-2007 | 11 | 470 | 5 | 7 | 493 | | | | | |
| B.1.1.33 | | RJ-2091 | 2 | 309 | 0 | 1 | 312 | | | | | |
| B.1.1.33 | | RJ-2342 | 0 | 111 | 0 | 0 | 111 | | | | | |

**Table S6.** Within-host diversity BR-UY-I28. The table shows the observed allele count for each synapomorphic site identified in clade BR-UY-I28

| | | | **1. 1471C>T (ORF1ab: R402R)** | | | | | **2. 1519C>T (ORF1ab: N418N)** | | | | |
|---|---|---|---|---|---|---|---|---|---|---|---|---|
| Lineage | Outbreak | Sample | A | C | G | T | num.reads | A | C | G | T | num.reads |
| B.1.1.28 | Bella Unión | M151 | 0 | 3 | 0 | 29 | 32 | 1 | 1 | 0 | 29 | 31 |

| | | | | | | | | | | | | |
|---|---|---|---|---|---|---|---|---|---|---|---|---|
| B.1.1.28 | Bella Unión | M152 | 0 | 1 | 0 | 14 | 15 | 0 | 1 | 0 | 15 | 16 |
| B.1.1.28 | Bella Unión | M163 | 1 | 4 | 2 | 43 | 50 | 3 | 1 | 1 | 45 | 50 |
| B.1.1.28 | Bella Unión | M157 | 2 | 6 | 0 | 171 | 179 | 0 | 9 | 3 | 163 | 175 |
| | | | **3. 5995A>G (ORF1ab: Q1910Q)** | | | | | **4. 9605T>C (ORF1ab: F3114L)** | | | | |
| Lineage | Outbreak | Sample | A | C | G | T | num.reads | A | C | G | T | num.reads |
| B.1.1.28 | Bella Unión | M151 | 23 | 0 | 171 | 1 | 195 | 1 | 201 | 0 | 14 | 216 |
| B.1.1.28 | Bella Unión | M152 | 8 | 0 | 30 | 0 | 38 | 0 | 49 | 0 | 2 | 51 |
| B.1.1.28 | Bella Unión | M163 | 1 | 0 | 7 | 0 | 8 | 0 | 4 | 0 | 0 | 4 |
| B.1.1.28 | Bella Unión | M157 | 8 | 0 | 34 | 0 | 42 | 0 | 10 | 0 | 1 | 11 |
| | | | **5. 9693C>T (ORF1ab: A3143V)** | | | | | **6. 15606T>C (ORF1ab: N5113N)** | | | | |
| Lineage | Outbreak | Sample | A | C | G | T | num.reads | A | C | G | T | num.reads |
| B.1.1.28 | Bella Unión | M151 | 0 | 6 | 0 | 213 | 219 | 0 | 20 | 0 | 1 | 21 |
| B.1.1.28 | Bella Unión | M152 | 1 | 1 | 0 | 49 | 51 | 0 | 15 | 0 | 0 | 15 |
| B.1.1.28 | Bella Unión | M163 | 0 | 0 | 0 | 4 | 4 | 1 | 288 | 0 | 18 | 307 |
| B.1.1.28 | Bella Unión | M157 | 0 | 0 | 0 | 10 | 10 | 0 | 722 | 0 | 44 | 766 |
| | | | **7. 23416A>T (Spike protein S1: T606T)** | | | | | **8. 26159T>C (ORF3a: V256A)** | | | | |
| Lineage | Outbreak | Sample | A | C | G | T | num.reads | A | C | G | T | num.reads |
| B.1.1.28 | Bella Unión | M151 | 2 | 0 | 5 | 65 | 72 | 6 | 884 | 0 | 53 | 943 |
| B.1.1.28 | Bella Unión | M152 | 0 | 0 | 0 | 2 | 2 | 0 | 299 | 0 | 27 | 326 |
| B.1.1.28 | Bella Unión | M163 | 37 | 27 | 42 | 845 | 951 | 0 | 59 | 0 | 3 | 62 |
| B.1.1.28 | Bella Unión | M157 | 49 | 27 | 36 | 843 | 955 | 0 | 287 | 0 | 13 | 300 |
| | | | **9. 28508A>T (N: S79C)** | | | | | **10. 29367C>T (N: P365L)** | | | | |
| Lineage | Outbreak | Sample | A | C | G | T | num.reads | A | C | G | T | num.reads |
| B.1.1.28 | Bella Unión | M151 | 17 | 7 | 2 | 453 | 479 | 0 | 59 | 2 | 554 | 615 |
| B.1.1.28 | Bella Unión | M152 | 2 | 1 | 0 | 98 | 101 | 0 | 24 | 0 | 195 | 219 |
| B.1.1.28 | Bella Unión | M163 | 14 | 6 | 1 | 642 | 663 | 3 | 32 | 1 | 391 | 427 |
| B.1.1.28 | Bella Unión | M157 | 18 | 6 | 4 | 519 | 547 | 0 | 20 | 0 | 217 | 237 |
| | | | **11. 29737G>T (3'-UTR)** | | | | | | | | | |
| Lineage | Outbreak | Sample | A | C | G | T | num.reads | | | | | |
| B.1.1.28 | Bella Unión | M151 | 2 | 6 | 30 | 567 | 605 | | | | | |
| B.1.1.28 | Bella Unión | M152 | 0 | 3 | 7 | 203 | 213 | | | | | |
| B.1.1.28 | Bella Unión | M163 | 0 | 0 | 2 | 46 | 48 | | | | | |
| B.1.1.28 | Bella Unión | M157 | 0 | 0 | 0 | 3 | 3 | | | | | |

**Table S7.** Within-host diversity BR-RS-I33. The table shows the observed allele count for each synapomorphic site identified in clade BR-RS-I33

| | **1. 15654C>T (ORF1ab: D5129D)** | | | | |
|---|---|---|---|---|---|
| Sample | A | C | G | T | num.reads |
| RS-15276 | 3 | 18 | 0 | 461 | 482 |
| RS-15287 | 4 | 8 | 4 | 470 | 486 |
| RS-6179 | 4 | 3 | 10 | 4130 | 4147 |
| RS-6180 | 0 | 5 | 4 | 4997 | 5006 |
| RS-6187 | 3 | 3 | 1 | 3193 | 3200 |
| RS-6190 | 7 | 8 | 6 | 5780 | 5801 |
| RS-6197 | 3 | 7 | 1 | 3805 | 3816 |
| RS-6198 | 1 | 8 | 0 | 3045 | 3054 |
| RS-6227 | 7 | 3 | 5 | 4325 | 4340 |
| RS-6228 | 2 | 1 | 4 | 3072 | 3079 |

| | | | | | |
|---|---|---|---|---|---|
| RS-6241 | 7 | 1 | 4 | 3997 | 4009 |
| RS-6242 | 7 | 11 | 2 | 6566 | 6586 |

**Table S8.** Linear model. Obtained coefficients for the explanatory variables "SEQ", "SAMPLE" and "MUTATION" are shown. For the calculations, the observations were filtered according to a minimum coverage of 100 reads. Allele frequencies and Shannon Entropy (H) were estimated. The linear model was implemented in R with lm(formula = He1.4 ~ SEQ + SAMPLE + MUTATION, data = tab.f). Obtained Residual standard error was 0.1026 on 614 degrees of freedom. Multiple R-squared: 0.7267, Adjusted R-squared: 0.6844. F-statistic: 17.19 on 95 and 614 DF and p-value: < 2.2e-16. Coefficients: 3 not defined because of singularities. The formatted data for statistical analysis is available upon request.

| Coefficient | Estimate | Std.Error | t value | Pr(>\|t\|) | |
|---|---|---|---|---|---|
| (Intercept) | 0.4303370 | 0.1026450 | 4192 | 3.17e-05 | *** |
| SEQIonTorrent | -0.0197866 | 0.1076802 | -184 | 0.854268 | |
| SEQONT | 0.3252601 | 0.1094966 | 2971 | 0.003089 | ** |
| SAMPLEM11 | -0.0327176 | 0.0384724 | -850 | 0.395425 | |
| SAMPLEM119 | -0.0405952 | 0.0468754 | -866 | 0.386815 | |
| SAMPLEM12 | -0.0325717 | 0.0384724 | -847 | 0.397534 | |
| SAMPLEM13 | -0.0203398 | 0.0384724 | -529 | 0.597214 | |
| SAMPLEM130 | -0.0504425 | 0.0468708 | -1076 | 0.282258 | |
| SAMPLEM132 | -0.0221382 | 0.0468708 | -472 | 0.636863 | |
| SAMPLEM133 | -0.0174062 | 0.0460311 | -378 | 0.705457 | |
| SAMPLEM135 | -0.0012343 | 0.0460311 | -27 | 0.978616 | |
| SAMPLEM136 | -0.0207418 | 0.0460311 | -451 | 0.652433 | |
| SAMPLEM14 | -0.0397404 | 0.0384724 | -1033 | 0.302030 | |
| SAMPLEM15 | -0.0323473 | 0.0384724 | -841 | 0.400791 | |
| SAMPLEM151 | 0.0391147 | 0.1094966 | 357 | 0.721048 | |
| SAMPLEM152 | 0.0178085 | 0.1313678 | 136 | 0.892212 | |
| SAMPLEM154 | -0.0464515 | 0.0479261 | -969 | 0.332811 | |
| SAMPLEM155 | -0.0353734 | 0.0468708 | -755 | 0.450719 | |
| SAMPLEM157 | 0.0011578 | 0.1339953 | 9 | 0.993108 | |
| SAMPLEM163 | -0.0012125 | 0.1385434 | -9 | 0.993020 | |
| SAMPLEM40 | -0.0674552 | 0.0460311 | -1465 | 0.143316 | |
| SAMPLEM43 | -0.0522903 | 0.0468708 | -1116 | 0.265018 | |
| SAMPLEM49 | -0.0428898 | 0.0460311 | -932 | 0.351828 | |
| SAMPLEM5 | -0.0222417 | 0.0384724 | -578 | 0.563394 | |
| SAMPLEM52 | -0.0338867 | 0.0491049 | -690 | 0.490400 | |
| SAMPLEM53 | -0.0265017 | 0.0548308 | -483 | 0.629029 | |
| SAMPLEM54 | -0.0667156 | 0.0491049 | -1359 | 0.174762 | |
| SAMPLEM57 | -0.0293009 | 0.0460311 | -637 | 0.524659 | |
| SAMPLEM59 | -0.0507807 | 0.0548308 | -926 | 0.354740 | |
| SAMPLEM60 | -0.0216759 | 0.0525024 | -413 | 0.679856 | |
| SAMPLEM62 | -0.0290056 | 0.0506147 | -573 | 0.566809 | |
| SAMPLEM63 | 0.0032653 | 0.0525024 | 62 | 0.950429 | |
| SAMPLEM64 | -0.0239212 | 0.0460311 | -520 | 0.603478 | |
| SAMPLEM65 | 0.0153287 | 0.0469325 | 327 | 0.744074 | |
| SAMPLEM66 | 0.0124419 | 0.0479299 | 260 | 0.795271 | |
| SAMPLEM67 | -0.0153023 | 0.0479299 | -319 | 0.749634 | |
| SAMPLEM68 | 0.0164567 | 0.0469325 | 351 | 0.725974 | |
| SAMPLEM69 | 0.0081191 | 0.0469325 | 173 | 0.862711 | |
| SAMPLEM70 | -0.0241049 | 0.0479299 | -503 | 0.615200 | |
| SAMPLEM71 | 0.0253492 | 0.0469325 | 540 | 0.589310 | |
| SAMPLEM72 | -0.0396576 | 0.0468708 | -846 | 0.397824 | |

| | | | | | |
|---|---|---|---|---|---|
| SAMPLEM73 | -0.0919521 | 0.0506220 | -1816 | 0.069790 | . |
| SAMPLEM79 | -0.0972276 | 0.0478620 | -2031 | 0.042643 | * |
| SAMPLEM8 | 0.0030301 | 0.0390998 | 77 | 0.938255 | |
| SAMPLEM85 | -0.0522872 | 0.0460311 | -1136 | 0.256437 | |
| SAMPLEM87 | -0.0082634 | 0.0460311 | -180 | 0.857591 | |
| SAMPLEM9 | -0.0162466 | 0.0419046 | -388 | 0.698369 | |
| SAMPLEM91 | -0.0347698 | 0.0460311 | -755 | 0.450326 | |
| SAMPLEM93 | -0.0453471 | 0.0460311 | -985 | 0.324943 | |
| SAMPLEM94 | -0.0351558 | 0.0460311 | -764 | 0.445316 | |
| SAMPLEM95 | -0.0388039 | 0.0460311 | -843 | 0.399561 | |
| SAMPLEM97 | -0.0211658 | 0.0460311 | -460 | 0.645812 | |
| SAMPLEM98 | -0.0535552 | 0.0460311 | -1163 | 0.245096 | |
| SAMPLERJ-2007 | -0.0576248 | 0.0460311 | -1252 | 0.211095 | |
| SAMPLERJ-2091 | 0.0617864 | 0.1076802 | 574 | 0.566317 | |
| SAMPLERJ-2342 | 0.0708355 | 0.1076802 | 658 | 0.510893 | |
| SAMPLERS-15273 | 0.0198147 | 0.1076802 | 184 | 0.854063 | |
| SAMPLERS-15276 | -0.0221979 | 0.1094966 | -203 | 0.839416 | |
| SAMPLERS-15280 | -0.0322250 | 0.0469308 | -687 | 0.492563 | |
| SAMPLERS-15287 | -0.0401016 | 0.1094966 | -366 | 0.714315 | |
| SAMPLERS-6177 | 0.1111980 | 0.1076802 | 1033 | 0.302166 | |
| SAMPLERS-6179 | 0.0296736 | 0.1451619 | 204 | 0.838095 | |
| SAMPLERS-6180 | -0.0473636 | 0.1451619 | -326 | 0.744323 | |
| SAMPLERS-6187 | -0.0263455 | 0.1451619 | -181 | 0.856043 | |
| SAMPLERS-6188 | 0.0840227 | 0.1098865 | 765 | 0.444785 | |
| SAMPLERS-6190 | 0.0190182 | 0.1451619 | 131 | 0.895807 | |
| SAMPLERS-6197 | -0.0045496 | 0.1451619 | -31 | 0.975007 | |
| SAMPLERS-6198 | -0.0084149 | 0.1451619 | -58 | 0.953792 | |
| SAMPLERS-6203 | 0.0610583 | 0.1092096 | 559 | 0.576302 | |
| SAMPLERS-6208 | 0.1998152 | 0.1086602 | 1839 | 0.066413 | . |
| SAMPLERS-6222 | 0.0402799 | 0.1076802 | 374 | 0.708481 | |
| SAMPLERS-6227 | 0.0143911 | 0.1451619 | 99 | 0.921061 | |
| SAMPLERS-6228 | -0.0234911 | 0.1451619 | -162 | 0.871496 | |
| SAMPLERS-6231 | 0.0621493 | 0.1080440 | 575 | 0.565352 | |
| SAMPLERS-6241 | -0.0008945 | 0.1451619 | -6 | 0.995085 | |
| SAMPLERS-6242 | NA | NA | NA | NA | |
| MUTATIONA2276G | -0.0517600 | 0.0197703 | -2618 | 0.009061 | ** |
| MUTATIONA23416T | 0.1563496 | 0.1454285 | 1075 | 0.282754 | |
| MUTATIONA28508T | -0.0300958 | 0.1244753 | -242 | 0.809031 | |
| MUTATIONA5089G | 0.0710363 | 0.0210444 | 3376 | 0.000783 | *** |
| MUTATIONA5995G | 0.0778204 | 0.1451619 | 536 | 0.592088 | |
| MUTATIONC12025T | -0.1550629 | 0.0193689 | -8006 | 5.96e-15 | *** |
| MUTATIONC1471T | -0.0092689 | 0.1644299 | -56 | 0.955065 | |
| MUTATIONC15108T | 0.0069632 | 0.0198457 | 351 | 0.725810 | |
| MUTATIONC1519T | 0.0521031 | 0.1644299 | 317 | 0.751449 | |
| MUTATIONC15654T | NA | NA | NA | NA | |
| MUTATIONC18252T | -0.0221625 | 0.0193604 | -1145 | 0.252765 | |
| MUTATIONC28093T | -0.0954197 | 0.0195788 | -4874 | 1.40e-06 | *** |
| MUTATIONC28775T | 0.0639506 | 0.0199810 | 3201 | 0.001442 | ** |
| MUTATIONC29367T | 0.0602654 | 0.1244753 | 484 | 0.628448 | |
| MUTATIONC8293T | -0.0568249 | 0.0202967 | -2800 | 0.005276 | ** |
| MUTATIONC9693T | -0.1304916 | 0.1451619 | -899 | 0.369038 | |
| MUTATIONG189T | -0.1314978 | 0.0196020 | -6708 | 4.47e-11 | *** |

| | | | | | |
|---|---|---|---|---|---|
| MUTATIONG23311T | -0.1833839 | 0.0190945 | -9604 | 2.00E-016 | *** |
| MUTATIONG24872T | -0.0518007 | 0.0190176 | -2724 | 0.006636 | ** |
| MUTATIONG29737T | -0.0091636 | 0.1308472 | -70 | 0.944190 | |
| MUTATIONT15606C | 0.0084781 | 0.1454285 | 58 | 0.953531 | |
| MUTATIONT26159C | -0.0068957 | 0.1260216 | -55 | 0.956381 | |
| MUTATIONT9605C | NA | NA | NA | NA | |

**Table S9.** GISAID acknowledgement table for B1.1.33 sequences

GISAID

We gratefully acknowledge the following Authors from the Originating laboratories responsible for obtaining the specimens, as well as the Submitting laboratories where the genome data were generated and shared via GISAID, on which this research is based.

All Submitters of data may be contacted directly via www.gisaid.org

| Accession ID | Originating Laboratory | Submitting Laboratory | Authors |
|---|---|---|---|
| EPI_ISL_418371 | Public Health Ontario Laboratories | Public Health Ontario Laboratories | Alireza Eshaghi, Samir N Patel, Jonathan B Gubbay, Vanessa G Allen, Christine Frantz, Aimin Li, Sandeep Nagra |
| EPI_ISL_425171 | University of Wisconsin-Madison AIDS Vaccine Research Laboratories | University of Wisconsin-Madison AIDS Vaccine Research Laboratories | Gage Moreno, Katarina Braun, et al. AIDS Vaccine Research Laboratories |
| EPI_ISL_425373 | Department of Pathology, University of Cambridge | COVID-19 Genomics UK (COG-UK) Consortium | Luke W Meredith, M. Estee Torok , Myra Hosmillo, William L. Hamilton, Martin D. Curran, Theresa Feltwell, Anna Yakovleva, Charlotte J. Houldcroft, Aminu S. Jahun, Sarah L. Caddy, Ian Goodfellow |
| EPI_ISL_426899 | Royal Darwin Hospital Pathology | Microbiological Diagnostic Unit Public Health Laboratory and Victorian Infectious Diseases Reference Laboratory, Doherty Institute | Meumann, E., Caly L., Seemann T., Sait, M., Schultz M., Druce J., Sherry, N. |
| EPI_ISL_427131 | Victorian Infectious Diseases Reference Laboratory (VIDRL) | Microbiological Diagnostic Unit Public Health Laboratory and Victorian Infectious Diseases Reference Laboratory, Doherty Institute | Caly L., Seemann T., Sait, M., Schultz M., Druce J., Sherry, N. |
| EPI_ISL_427294, EPI_ISL_427295, EPI_ISL_427296, EPI_ISL_427297, EPI_ISL_427298, EPI_ISL_427302, EPI_ISL_427303, EPI_ISL_427304 | Instituto Oswaldo Cruz FIOCRUZ - Laboratory of Respiratory Viruses and Measles (LVRS) | Instituto Oswaldo Cruz FIOCRUZ - Laboratory of Respiratory Viruses and Measles (LVRS) | Paola Resende, Fernando Motta, Luciana Appolinario, Sunando Roy, Aline Mattos, Milene Miranda, Cristiana Garcia, Braulia Caetano, Maria Ogrzewalska, Priscila Born, Jonathan Lopes, Marilda Siqueira |
| EPI_ISL_428294 | University of Wisconsin-Madison AIDS Vaccine Research Laboratories | University of Wisconsin-Madison AIDS Vaccine Research Laboratories | Gage Moreno, Katarina Braun, et al. AIDS Vaccine Research Laboratories |
| EPI_ISL_430814, EPI_ISL_430815, EPI_ISL_430817 | Laboratorio de Virología del Hospital de Niños Dr. Ricardo Gutierrez | Área de Secuenciación del Laboratorio de Virología del Hospital de Niños Dr. Ricardo Gutierrez | Nabaes Jodar, MS; Goya, S; Natale, MI; Lusso, S; Gravis, E; Mistchenko, AS; Valinotto, LE; Viegas, M. |
| EPI_ISL_434803 | Houston Methodist Hospital | Houston Methodist Hospital | S. Wesley Long, Randall J. Olsen, Paul A. Christensen, David W. Bernard, James J. Davis, Maulik Shukla, Marcus Nguyen, Matthew Ojeda Saavedra, Concepcion C. Cantu, Prasanti Yerramilli, Layne Pruitt, Sishir Subedi, Heather Hendrickson, Ghazaleh Eskandari, Muthiah Kumaraswami, Jason S. McLellan, Hakon Jonsson, Kari Stefansson, and James M. Musser |
| EPI_ISL_436853, EPI_ISL_437100 | Michigan Department of Health and Human Services, Bureau of Laboratories | Michigan Department of Health and Human Services, Bureau of Laboratories | Blankenship HM, Riner D, Soehnlen MK |
| EPI_ISL_437806 | UW Virology Lab | UW Virology Lab | Pavitra Roychoudhury, Hong Xie, Keith Jerome, Alexander Greninger |
| EPI_ISL_444626 | NYU Langone Health | Departments of Pathology and Medicine, New York University School of Medicine | Maria Aguero-Rosenfeld, Brendan Belovarac, Margaret Black, Ludovic Boytard, John Cadley, Paolo Cotzia, John Chen, Dacia Dimartino, Xiaojun Feng, Tatyana Gindin, Emily Guzman, Adriana Heguy, Megan Hogan, Emily Huang, George Jour, Alireza Khodadadi-Jamayran, Lawrence H. Lin, Raven Luther, Andrew Lytle, Christian Marier, Matthew T. Maurano, Mark J. Mulligan, Peter Meyn, Raquel Ordonez Ciriza, Iman Osman, Jared Pinnell, Vanessa Raabe, Sitharam Ramaswami, Amy Rapkiewicz, Andre M. Ribeiro-dos-Santos, Marie Samanovic-Golden, Antonio Serrano, Guomiao Shen, Matija Snuderl, Theodore Vougiouklakis, Nick Vulpescu, Gael Westby, Paul Zappile, Yutong Zhang |
| EPI_ISL_445349 | HOSPITAL SAN JUAN DE DIOS | Instituto de Salud Publica de Chile | Andrés E Castillo, Bárbara Parra,Paz Tapia, Jaime Lagos, Loredana Arata, Alejandra Acevedo, Winston Andrade, Gabriel Leal, Carolina Tambley, Patricia Bustos, Rodrigo Fasce, Jorge Fernandez |
| EPI_ISL_445352 | HOSPITAL DEL PROFESOR | Instituto de Salud Publica de Chile | Andrés E Castillo, Bárbara Parra,Paz Tapia, Jaime Lagos, Loredana Arata, Alejandra Acevedo, Winston Andrade, Gabriel Leal, Carolina Tambley, Patricia Bustos, Rodrigo Fasce, Jorge Fernandez |
| EPI_ISL_445362 | BUPA SERVICIOS CLINICOS S.A | Instituto de Salud Publica de Chile | Andrés E Castillo, Bárbara Parra,Paz Tapia, Jaime Lagos, Loredana Arata, Alejandra Acevedo, Winston Andrade, Gabriel Leal, Carolina Tambley, Patricia Bustos, Rodrigo Fasce, Jorge Fernandez |
| EPI_ISL_445367 | ASISTENCIA PUBLICA DR.ALEJANDRO DEL RIO | Instituto de Salud Publica de Chile | Andrés E Castillo, Bárbara Parra,Paz Tapia, Jaime Lagos, Loredana Arata, Alejandra Acevedo, Winston Andrade, Gabriel Leal, Carolina Tambley, Patricia Bustos, Rodrigo Fasce, Jorge Fernandez |
| EPI_ISL_445369, EPI_ISL_445370 | HOSPITAL DE CARABINEROS | Instituto de Salud Publica de Chile | Andrés E Castillo, Bárbara Parra,Paz Tapia, Jaime Lagos, Loredana Arata, Alejandra Acevedo, Winston Andrade, Gabriel Leal, Carolina Tambley, Patricia Bustos, Rodrigo Fasce, Jorge Fernandez |
| EPI_ISL_445373 | HOSPITAL SAN JUAN DE DIOS | Instituto de Salud Publica de Chile | Andrés E Castillo, Bárbara Parra,Paz Tapia, Jaime Lagos, Loredana Arata, Alejandra Acevedo, Winston Andrade, Gabriel Leal, Carolina Tambley, Patricia Bustos, Rodrigo Fasce, Jorge Fernandez |
| EPI_ISL_450873 | Evandro Chagas Institute | Evandro Chagas Institute | Santos, M.C.; Silva, A.M.; Junior, W.D.C.; Barbagelata, L.S.; Ferreira, J.A.; Sousa, E.M.A.; da Silva, P.S.; Martins, L.C.; Sousa Junior, E.C.; Viana, G.M.R |
| EPI_ISL_450874 | Evandro Chagas Institute | Evandro Chagas Institute | Santos, M.C.; Silva, A.M.; Junior, W.D.C.; Barbagelata, L.S.; Ferreira, J.A.; Sousa, E.M.A.; da Silva, P.S.; Martins, L.C.;Sousa Junior, E.C.;Viana, G.M.R |
| EPI_ISL_451558 | Medlab Pathology | NSW Health Pathology - Institute of Clinical Pathology and Medical Research; Westmead Hospital; University of Sydney | CIDM-PH et al. |
| EPI_ISL_451594 | Childrens Hospital Westmead | NSW Health Pathology - Institute of Clinical Pathology and Medical Research; Westmead Hospital; University of Sydney | CIDM-PH et al. |
| EPI_ISL_452318 | Michigan Department of Health and Human Services, Bureau of Laboratories | Michigan Department of Health and Human Services, Bureau of Laboratories | Blankenship HM, Riner D, Soehnlen MK |
| EPI_ISL_454057, EPI_ISL_454172, EPI_ISL_454311 | unknown | Instituto Nacional de Saude (INSA) | Borges et al |
| EPI_ISL_454376 | UPMC Clinical Microbiology Laboratory | Microbial Genome Sequencing Center, Microbial Genomic Epidemiological Laboratory | Mustapha M. Mustapha, Jane W. Marsh, Dan Snyder, Marissa P. Griffith, Stephanie L. Mitchell, Vatsala R. Srinivasa, Kady D. Waggle, Chinelo Ezeonwuku, Vaughn S. Cooper, Lee H. Harrison |
| EPI_ISL_456071, EPI_ISL_456072, EPI_ISL_456073, EPI_ISL_456074, EPI_ISL_456075 | Laboratory of Respiratory Viruses and Measles, Oswaldo Cruz Institute, FIOCRUZ | Laboratory of Respiratory Viruses and Measles, Oswaldo Cruz Institute, FIOCRUZ | Paola Resende, Luciana Appolinario, Fernando Motta, Aline Mattos, Milene Miranda, Cristiana Garcia, Braulia Caetano, Maria Ogrzewalska, Jonathan Lopes, Marilda Siqueira |
| EPI_ISL_456076, EPI_ISL_456077 | LACEN RJ - Laboratório Central de Saúde Pública Noel Nutels | Laboratory of Respiratory Viruses and Measles, Oswaldo Cruz Institute, FIOCRUZ | Paola Resende, Luciana Appolinario, Fernando Motta, Aline Mattos, Milene Miranda, Cristiana Garcia, Braulia Caetano, Maria Ogrzewalska, Jonathan Lopes, Marilda Siqueira |
| EPI_ISL_456079, EPI_ISL_456080, EPI_ISL_456081 | Laboratory of Respiratory Viruses and Measles, Oswaldo Cruz Institute, FIOCRUZ | Laboratory of Respiratory Viruses and Measles, Oswaldo Cruz Institute, FIOCRUZ | Paola Resende, Luciana Appolinario, Fernando Motta, Aline Mattos, Milene Miranda, Cristiana Garcia, Braulia Caetano, Maria Ogrzewalska, Jonathan Lopes, Marilda Siqueira |
| EPI_ISL_456082, EPI_ISL_456083 | LACEN RJ - Laboratório Central de Saúde Pública Noel Nutels | Laboratory of Respiratory Viruses and Measles, Oswaldo Cruz Institute, FIOCRUZ | Paola Resende, Luciana Appolinario, Fernando Motta, Aline Mattos, Milene Miranda, Cristiana Garcia, Braulia Caetano, Maria Ogrzewalska, Jonathan Lopes, Marilda Siqueira |
| EPI_ISL_456084, EPI_ISL_456085, EPI_ISL_456086, EPI_ISL_456087, EPI_ISL_456089, EPI_ISL_456090, EPI_ISL_456091, EPI_ISL_456092, EPI_ISL_456093, EPI_ISL_456094, EPI_ISL_456095, EPI_ISL_456096, EPI_ISL_456097, EPI_ISL_456098, EPI_ISL_456099, EPI_ISL_456100, EPI_ISL_456101, EPI_ISL_456102, EPI_ISL_456103, EPI_ISL_456104, EPI_ISL_456105, EPI_ISL_456106 | | | |
| see above | Laboratory of Respiratory Viruses and Measles, Oswaldo Cruz Institute, FIOCRUZ | Laboratory of Respiratory Viruses and Measles, Oswaldo Cruz Institute, FIOCRUZ | Paola Resende, Luciana Appolinario, Fernando Motta, Aline Mattos, Milene Miranda, Cristiana Garcia, Braulia Caetano, Maria Ogrzewalska, Jonathan Lopes, Marilda Siqueira |
| EPI_ISL_457796 | Johns Hopkins Hospital Department of Pathology | Johns Hopkins Hospital Department of Pathology | Peter M. Thielen, Thomas Mehoke, Shirlee Wohl, Srividya Ramakrishnan, Melanie Kirsche, Amanda Ernlund, Craig Howser, Kristina Zudock, Oluwaseun Falade-Nwulia, Norah Sadowski, Paul Morris, Mark Hopkins, Yunfan Fan, Nidia Trovao, Victoria Gniazdowski, Michael C. Schatz, Stuart C. Ray, Winston Timp, Heba H. Mostafa |
| EPI_ISL_457953 | Laboratorio de Biología Molecular Asociación Española Primera en Salud | Departments of Pathology and Medicine, New York University School of Medicine | Maria Victoria Elizondo, Maria Noel Zubillaga, Gonzalo Manrique, Paul Zappile, Gael Westby, Matthew T Maurano, Christian Marier, Adriana Heguy |
| EPI_ISL_458138, EPI_ISL_458139, EPI_ISL_458142, EPI_ISL_458143, EPI_ISL_458144, EPI_ISL_458145, EPI_ISL_458148, EPI_ISL_458149 | Evandro Chagas Institute | Evandro Chagas Institute | Santos, M.C.; Silva, A.M.; Junior, W.D.C.; Barbagelata, L.S.; Ferreira, J.A.; Sousa, E.M.A.; da Silva, P.S.; Resque, H.R; Martins, L.C.; Sousa Junior, E.C.;Viana, G.M.R |
| EPI_ISL_460134, EPI_ISL_460202, EPI_ISL_460215, EPI_ISL_460236, EPI_ISL_460403 | Massachusetts General Hospital | Infectious Disease Program, Broad Institute of Harvard and MIT | Lemieux,J.E., Siddle,K.J., Shaw,B., Adams,G., Pierce,V., Turbett,S., Anahtar,M., Branda,J., Slater,D., Harris,J., Lin,A.E., Gladden-Young,A., Lagerborg,K., Rudy,M., DeRuff,K., Carter,A., Normandin,E., Bauer,M., Reilly,S., Tomkins-Tinch,C., Loreth,C., Chaluvadi,S., Neumann,A., Cusick,C., Chapman,S.B., Gnirke,A., Flowers,K., Cerrato,F., Birren,B.W., Gallagher,G., Smole,S., Park,D.J., MacInnis,B.L., Ryan,E., LaRocque,R., Rosenberg,E., Sabeti,P.C. |
| EPI_ISL_465088 | Respiratory Virus Unit, Microbiology Services Colindale, Public Health England | Respiratory Virus Unit, Microbiology Services Colindale, Public Health England | PHE Covid Sequencing Team |
| EPI_ISL_467345, EPI_ISL_467347, EPI_ISL_467348, EPI_ISL_467349, EPI_ISL_467350, EPI_ISL_467351, EPI_ISL_467352, EPI_ISL_467353, EPI_ISL_467355, EPI_ISL_467357, EPI_ISL_467358, EPI_ISL_467360, EPI_ISL_467361, EPI_ISL_467362, EPI_ISL_467363, EPI_ISL_467364, EPI_ISL_467365, EPI_ISL_467367, EPI_ISL_467368, EPI_ISL_467369, EPI_ISL_467370, EPI_ISL_467371 | | | |
| see above | Laboratory of Respiratory Viruses and Measles, Oswaldo Cruz Institute, FIOCRUZ | Laboratory of Respiratory Viruses and Measles, Oswaldo Cruz Institute, FIOCRUZ | Paola Resende, Luciana Appolinario, Fernando Motta, Anna Carolina Paixão, Ana Carolina Mendonça, Aline Mattos, Milene Miranda, Cristiana Garcia, Braulia Caetano, Maria Ogrzewalska, Jonathan Lopes, Marilda Siqueira |

**Table S10.** GISAID acknowledgement table for B1.1.28 sequences

We gratefully acknowledge the following Authors from the Originating laboratories responsible for obtaining the specimens, as well as the Submitting laboratories where the genome data were generated and shared via GISAID, on which this research is based.

All Submitters of data may be contacted directly via www.gisaid.org

| Accession ID | Originating Laboratory | Submitting Laboratory | Authors |
|---|---|---|---|
| EPI_ISL_416036 | National Influenza Center - Instituto Adolfo Lutz | Instituto Adolfo Lutz, Interdiciplinary Procedures Center, Strategic Laboratory | Claudio Tavares Sacchi, Claudia Regina Gonçalves, Carlos Henrique Camargo, Erica Valessa Ramos Gomes, Fabiana Cristina Pereira dos Santos, Daniela Bernardes Borges da Silva, Simone Guadagnucci Morillo, Adriano Abbud, Adriana Bugno, Maria do Carmo Sampaio Tavares Timenetsky, Terezinha Maria de Paiva |
| EPI_ISL_427292 | LACEN-AL - Laboratorio Central de Alagoas | Instituto Oswaldo Cruz FIOCRUZ - Laboratory of Respiratory Viruses and Measles (LVRS) | Paola Resende, Fernando Motta, Luciana Appolinario, Sunando Roy, Aline Mattos, Milene Miranda, Cristiana Garcia, Braulia Caetano, Maria Ogrzewalska, Priscila Born, Jonathan Lopes, Marilda Siqueira |
| EPI_ISL_431180, EPI_ISL_431240 | Fujian Center for Disease Control and Prevention | Fujian Center for Disease Control and Prevention | Lin Qi, Huang Zhimiao, Zhang Yanhua, Weng Yuwei |
| EPI_ISL_445380 | Ramathibodi Hospital | COVID-19 Network Investigations (CONI) Alliance | Elizabeth Batty, Wasun Chantratita, Thanat Chookajorn, Stefan Fernandez, Angkana Huang, Anthony R. Jones, Khajohn Joonsalak, Chonticha Klungtong, Theerarat Kochakarn, Namfon Kotanan, Krittikorn Kumpornsin, Wudtichai Manasatienkij, Bhakbhoom Panthan, Ekawat Pasomsub, Kingkan Rakmanee, Insee Sensorn, Janjira Thaipadungpanit, Arporn Wangwiwatsin, Treewat Watthanachockchai |
| EPI_ISL_456088 | LACEN RJ - Laboratório Central de Saúde Pública Noel Nutels | Laboratory of Respiratory Viruses and Measles, Oswaldo Cruz Institute, FIOCRUZ | Paola Resende, Luciana Appolinario, Fernando Motta, Aline Mattos, Milene Miranda, Cristiana Garcia, Braulia Caetano, Maria Ogrzewalska, Jonathan Lopes, Marilda Siqueira |
| EPI_ISL_456869 | West of Scotland Specialist Virology Centre, NHSGGC / MRC-University of Glasgow Centre for Virus Research | COVID-19 Genomics UK (COG-UK) Consortium | Ana da Silva Filipe, Natasha Johnson, Kathy Smollett, Daniel Mair, Stephen Carmichael, Lily Tong, Jenna Nichols, Elihu Aranday-Cortes, Kirstyn Brunker, Yasmin Parr, Kyriaki Nomikou; Sarah McDonald, Marc Niebel, Patawee Asamaphan; Richard Orton, Joseph Hughes, Sreenu Vattipally, David L Robertson; Alasdair MacLean, Rory Gunson; Kathy Li, Natasha Jesudason, Rajiv Shah, James Shepherd, Antonia Ho, Emma Thomson |
| EPI_ISL_458140, EPI_ISL_458141, EPI_ISL_458146, EPI_ISL_458147 | Evandro Chagas Institute | Evandro Chagas Institute | Santos, M.C.; Silva, A.M.; Junior, W.D.C.; Barbagelata, L.S.; Ferreira, J.A.; Sousa, E.M.A.; da Silva, P.S.; Resque, H.R; Martins, L.C.; Sousa Junior, E.C.;Viana, G.M.R |
| EPI_ISL_461606, EPI_ISL_461678 | West of Scotland Specialist Virology Centre, NHSGGC / MRC-University of Glasgow Centre for Virus Research | COVID-19 Genomics UK (COG-UK) Consortium | Ana da Silva Filipe, Natasha Johnson, Kathy Smollett, Daniel Mair, Stephen Carmichael, Lily Tong, Jenna Nichols, Elihu Aranday-Cortes, Kirstyn Brunker, Yasmin Parr, Kyriaki Nomikou; Sarah McDonald, Marc Niebel, Patawee Asamaphan; Richard Orton, Joseph Hughes, Sreenu Vattipally, David L Robertson; Alasdair MacLean, Rory Gunson; Kathy Li, Natasha Jesudason, Rajiv Shah, James Shepherd, Antonia Ho, Emma Thomson |
| EPI_ISL_467356, EPI_ISL_467359, EPI_ISL_467366 | Laboratory of Respiratory Viruses and Measles, Oswaldo Cruz Institute, FIOCRUZ | Laboratory of Respiratory Viruses and Measles, Oswaldo Cruz Institute, FIOCRUZ | Paola Resende, Luciana Appolinario, Fernando Motta, Anna Carolina Paixão, Ana Carolina Mendonça, Aline Mattos, Milene Miranda, Cristiana Garcia, Braulia Caetano, Maria Ogrzewalska, Jonathan Lopes, Marilda Siqueira |
| EPI_ISL_468305, EPI_ISL_468307 | Centro de Vigilancia a Saude de Diadema | Instituto Adolfo Lutz, Interdiciplinary Procedures Center, Strategic Laboratory | Claudio Tavares Sacchi, Claudia Regina Gonçalves, Erica Valessa Ramos Gomes |
| EPI_ISL_468308 | Hospital Municipal do Tatuape Carmino Caricchio | Instituto Adolfo Lutz, Interdiciplinary Procedures Center, Strategic Laboratory | Claudio Tavares Sacchi, Claudia Regina Gonçalves, Erica Valessa Ramos Gomes |
| EPI_ISL_468311, EPI_ISL_468312 | Hospital Municipal Dr Ignacio Proenca de Gouvea | Instituto Adolfo Lutz, Interdiciplinary Procedures Center, Strategic Laboratory | Claudio Tavares Sacchi, Claudia Regina Gonçalves, Erica Valessa Ramos Gomes |
| EPI_ISL_468313 | Vigilancia Epidemiologica de São Bernardo do Campo | Instituto Adolfo Lutz, Interdiciplinary Procedures Center, Strategic Laboratory | Claudio Tavares Sacchi, Claudia Regina Gonçalves, Erica Valessa Ramos Gomes |
| EPI_ISL_468314 | CTA Centro de Testagem e Aconselhamento | Instituto Adolfo Lutz, Interdiciplinary Procedures Center, Strategic Laboratory | Claudio Tavares Sacchi, Claudia Regina Gonçalves, Erica Valessa Ramos Gomes |
| EPI_ISL_468315 | Hospital Municipal do Tatuape Carmino Caricchio | Instituto Adolfo Lutz, Interdiciplinary Procedures Center, Strategic Laboratory | Claudio Tavares Sacchi, Claudia Regina Gonçalves, Erica Valessa Ramos Gomes |
| EPI_ISL_468316 | UPA Vila Assis | Instituto Adolfo Lutz, Interdiciplinary Procedures Center, Strategic Laboratory | Claudio Tavares Sacchi, Claudia Regina Gonçalves, Erica Valessa Ramos Gomes |
| EPI_ISL_468318 | Hospital Universitario da USP | Instituto Adolfo Lutz, Interdiciplinary Procedures Center, Strategic Laboratory | Claudio Tavares Sacchi, Claudia Regina Gonçalves, Erica Valessa Ramos Gomes |
| EPI_ISL_468319 | Vigilancia Epidemiologica de São Bernardo do Campo | Instituto Adolfo Lutz, Interdiciplinary Procedures Center, Strategic Laboratory | Claudio Tavares Sacchi, Claudia Regina Gonçalves, Erica Valessa Ramos Gomes |
| EPI_ISL_468321 | Hospital Universitario da USP | Instituto Adolfo Lutz, Interdiciplinary Procedures Center, Strategic Laboratory | Claudio Tavares Sacchi, Claudia Regina Gonçalves, Erica Valessa Ramos Gomes |
| EPI_ISL_470600, EPI_ISL_470602, EPI_ISL_470604, EPI_ISL_470605, EPI_ISL_470608, EPI_ISL_470610, EPI_ISL_470612 | Hermes Pardini | Bioinformatics Laboratory / LNCC | Alexandra Gerber, Ana Paula Guimarães, Luiz Gonzaga Paula de Almeida, Ronaldo da Silva Francisco Junior, Mariane Talon, Filipe Romero, Átila Duque Rossi, Terezinha Marta Pereira, working group UFRJ, Jaqueline Goes de Jesus, Ingra Morales Claro, Ester Cerdeira Sabino, Nuno Rodrigues Faria, CADDE-group, Laboratorio Hermes Pardini, Laboratorio Simile, working group UFMG, Amilcar Tanuri, Carolina Voloch, Renato Santana Aguiar e Ana Tereza Vasconcelos |
| EPI_ISL_470638 | Laboratorio de Virologia Molecular / UFRJ | Bioinformatics Laboratory / LNCC | Alexandra Gerber, Ana Paula Guimarães, Luiz Gonzaga Paula de Almeida, Ronaldo da Silva Francisco Junior, Mariane Talon, Filipe Romero, Átila Duque Rossi, Terezinha Marta Pereira, working group UFRJ, Jaqueline Goes de Jesus, Ingra Morales Claro, Ester Cerdeira Sabino, Nuno Rodrigues Faria, CADDE-group, Laboratorio Hermes Pardini, Laboratorio Simile, working group UFMG, Amilcar Tanuri, Carolina Voloch, Renato Santana Aguiar e Ana Tereza Vasconcelos |
| EPI_ISL_470651, EPI_ISL_470653, EPI_ISL_470654 | Hermes Pardini | Bioinformatics Laboratory / LNCC | Alexandra Gerber, Ana Paula Guimarães, Luiz Gonzaga Paula de Almeida, Ronaldo da Silva Francisco Junior, Mariane Talon, Filipe Romero, Átila Duque Rossi, Terezinha Marta Pereira, working group UFRJ, Jaqueline Goes de Jesus, Ingra Morales Claro, Ester Cerdeira Sabino, Nuno Rodrigues Faria, CADDE-group, Laboratorio Hermes Pardini, Laboratorio Simile, working group UFMG, Amilcar Tanuri, Carolina Voloch, Renato Santana Aguiar e Ana Tereza Vasconcelos |
| EPI_ISL_471539 | Hospital Universitario da USP Sao Paulo | Instituto Adolfo Lutz, Interdiciplinary Procedures Center, Strategic Laboratory | Claudio Tavares Sacchi, Claudia Regina Gonçalves, Erica Valessa Ramos Gomes |
| EPI_ISL_471541 | Hospital Geral Santa Marcelina | Instituto Adolfo Lutz, Interdiciplinary Procedures Center, Strategic Laboratory | Claudio Tavares Sacchi, Claudia Regina Gonçalves, Erica Valessa Ramos Gomes |
| EPI_ISL_471545 | Hospital Sao Paulo de Ensino da Unifesp | Instituto Adolfo Lutz, Interdiciplinary Procedures Center, Strategic Laboratory | Claudio Tavares Sacchi, Claudia Regina Gonçalves, Erica Valessa Ramos Gomes |
| EPI_ISL_471546 | AMA DR Jose Soares Hungria | Instituto Adolfo Lutz, Interdiciplinary Procedures Center, Strategic Laboratory | Claudio Tavares Sacchi, Claudia Regina Gonçalves, Erica Valessa Ramos Gomes |
| EPI_ISL_471548 | Hospital do Servidor Público Estadual Francisco Morato de Oliveira | Instituto Adolfo Lutz, Interdiciplinary Procedures Center, Strategic Laboratory | Claudio Tavares Sacchi, Claudia Regina Gonçalves, Erica Valessa Ramos Gomes |
| EPI_ISL_471549 | Hospital Municipal Carmen Prudente | Instituto Adolfo Lutz, Interdiciplinary Procedures Center, Strategic Laboratory | Claudio Tavares Sacchi, Claudia Regina Gonçalves, Erica Valessa Ramos Gomes |
| EPI_ISL_471552 | Hospital Sancta Maggiore | Instituto Adolfo Lutz, Interdiciplinary Procedures Center, Strategic Laboratory | Claudio Tavares Sacchi, Claudia Regina Gonçalves, Erica Valessa Ramos Gomes |
| EPI_ISL_471556 | Pronto Socorro Jose Ibrahin | Instituto Adolfo Lutz, Interdiciplinary Procedures Center, Strategic Laboratory | Claudio Tavares Sacchi, Claudia Regina Gonçalves, Erica Valessa Ramos Gomes |
| EPI_ISL_471562, EPI_ISL_471581, EPI_ISL_471582 | Hosp. Municipal Prof. Dr. Alípio Corrêa Netto | Instituto Adolfo Lutz, Interdiciplinary Procedures Center, Strategic Laboratory | Claudio Tavares Sacchi, Claudia Regina Gonçalves, Erica Valessa Ramos Gomes |
| EPI_ISL_471647 | Hospital Municipal de Barueri Dr. Francisco Moran | Instituto Adolfo Lutz, Interdiciplinary Procedures Center, Strategic Laboratory | Claudio Tavares Sacchi, Claudia Regina Gonçalves, Erica Valessa Ramos Gomes |
| EPI_ISL_471648 | UBS e Pronto Socorro Jd. Jacira | Instituto Adolfo Lutz, Interdiciplinary Procedures Center, Strategic Laboratory | Claudio Tavares Sacchi, Claudia Regina Gonçalves, Erica Valessa Ramos Gomes |
| EPI_ISL_473651 | West of Scotland Specialist Virology Centre, NHSGGC / MRC-University of Glasgow Centre for Virus Research | COVID-19 Genomics UK (COG-UK) Consortium | Ana da Silva Filipe, Natasha Johnson, Kathy Smollett, Daniel Mair, Stephen Carmichael, Lily Tong, Jenna Nichols, Elihu Aranday-Cortes, Kirstyn Brunker, Yasmin Parr, Alice Broos, Kyriaki Nomikou; Sarah McDonald, Marc Niebel, Patawee Asamaphan; Richard Orton, Joseph Hughes, Sreenu Vattipally, David L Robertson; Alasdair MacLean, Rory Gunson; Kathy Li, Natasha Jesudason, Rajiv Shah, James Shepherd, Antonia Ho, Emma Thomson |
| EPI_ISL_476152, EPI_ISL_476156, EPI_ISL_476157, EPI_ISL_476159, EPI_ISL_476161, EPI_ISL_476162, EPI_ISL_476163, EPI_ISL_476165, EPI_ISL_476167, EPI_ISL_476169 | Laboratório de Patologia Clínica - UNICAMP | Laboratório de Estudos de Vírus Emergentes - UNICAMP | José Luiz Proença-Modena, Magnun Nueldo Nunes dos Santos, Angelica Schreiber, Julia Forato,Camila Simeoni, Marcilio Jorge Fumagalli, Mariene Ribeiro Amorim, Darlan da Silva Candido, Nuno Rodrigues Faria, Julien Theze, Luiz Gonzaga,Jaqueline Goes Jesus e William Marciel de Souza |
| EPI_ISL_476244, EPI_ISL_476259 | Hospital da Clínicas da Faculdade de Medicina da Universidade de São Paulo | Instituto de Medicina Tropical da Univesidade de São Paulo | Samples: Ingra Morales Claro, Erika Regina Manuli, Cecilia Salete Alencar, Carolina S. Lazar, Silvia F. Costa; Sequencing: Ingra Morales Claro, Jaqueline Goes de Jesus, Erika Regina Manuli, Flavia Cristina da Silva Sales, Thais de Moura Coletti, Camila Alves Maia da Silva, Mariana Severo Ramundo, Giula Magalhaes Ferreira, Darlan da Silva Candido, Julien Theze, Nuno Faria, Ester Sabino |
| EPI_ISL_476293, EPI_ISL_476312, EPI_ISL_476318, EPI_ISL_476321, EPI_ISL_476322 | DB Diagnósticos do Brasil | Instituto de Medicina Tropical da Univesidade de São Paulo | Samples: Nelson Gaburo Jr; Sequencing: Ingra Morales Claro, Jaqueline Goes de Jesus, Erika Regina Manuli, Flavia Cristina da Silva Sales, Thais de Moura Coletti, Camila Alves Maia da Silva, Mariana Severo Ramundo, Giula Magalhaes Ferreira, Darlan da Silva Candido, Julien Theze, Nuno Faria, Ester Sabino |
| EPI_ISL_476337, EPI_ISL_476338, | Laboratório de Patologia Clínica - UNICAMP | Laboratório de Estudos de Vírus | José Luiz Proença-Modena, Magnun Nueldo Nunes dos Santos, Angelica Schreiber, Julia Forato,Camila Simeoni, Marcilio Jorge Fumagalli, Mariene Ribeiro Amorim, Darlan da Silva Candido, Nuno Rodrigues Faria, Julien Theze, Luiz Gonzaga,Jaqueline Goes |

#### Section 3. Supplementary figures

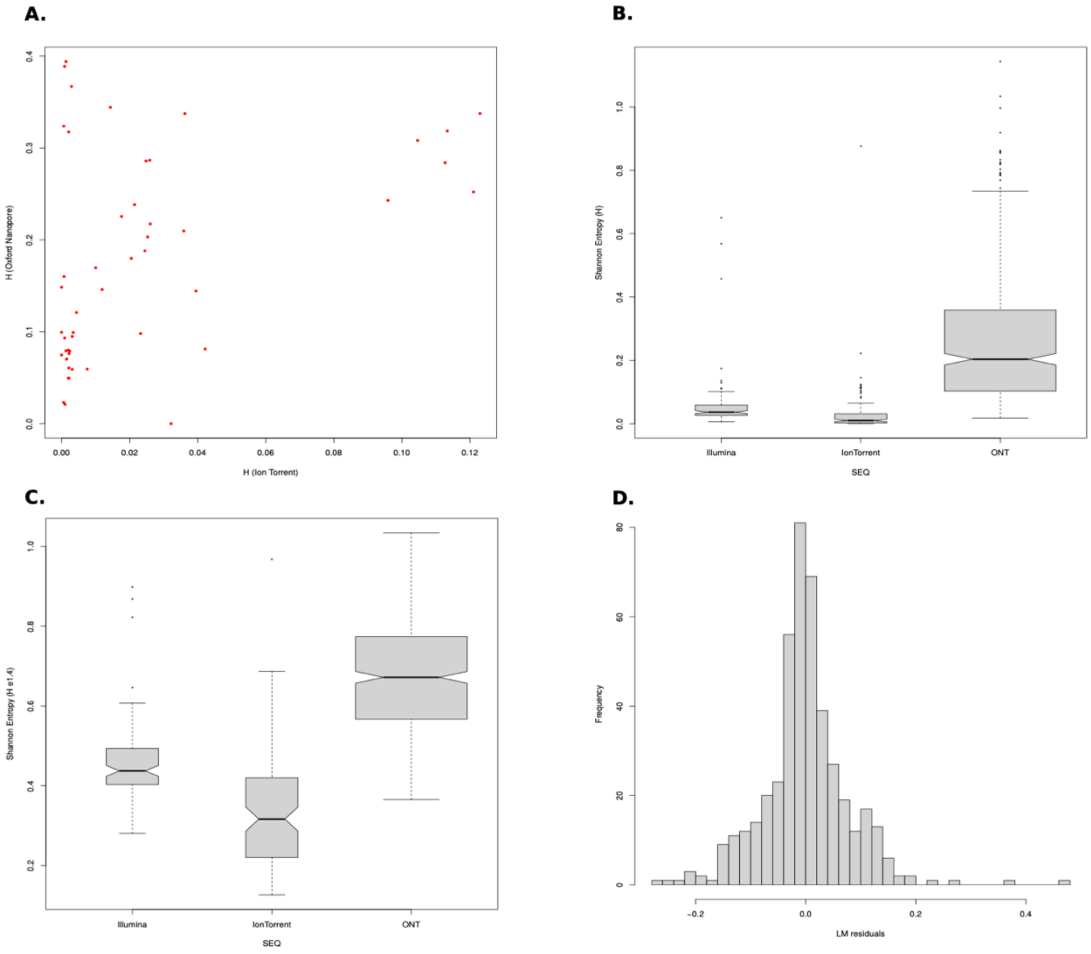

**Figure S1.** Effect of sequencing technology on intra-host diversity estimations. **A)** The samples were sequenced using three different technologies: Illumina, Ion Torrent, and ONT. In particular, samples M5, M8, M11, M12, M13, M14, and M15 were sequenced with both Ion Torrent and ONT technologies. The graphic depicts the Shannon Entropy values (H) obtained from allele frequencies obtained in each condition. As expected, due to the more error-prone ONT technology, H values are higher for data sequenced with ONT. **B)** After filtering those observations with less than 100 reads of genome coverage, the data consisted of 65, 63, and 264 points obtained by Illumina, Ion Torrent, and ONT sequencing, respectively. The boxplots show the distribution of H for each sequencing technology, H being higher for ONT. **C)** To better fit a normal distribution, H was square-root transformed (H exp ¼). **D)** The histogram depicts the distribution of residuals from the linear model implemented (H exp ¼ ~ SEQ + SAMPLE + MUTATION).

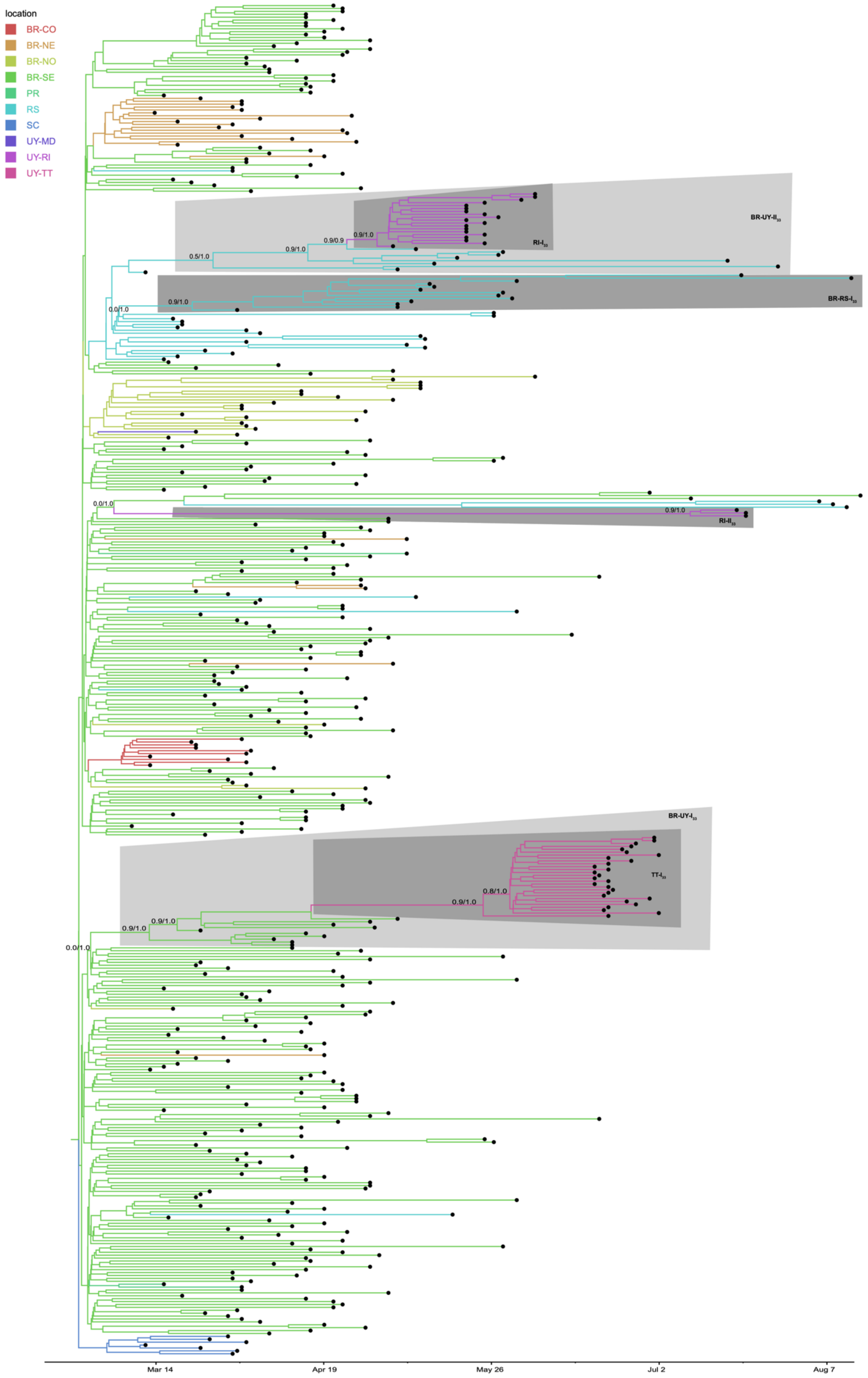

location
BR-CO
BR-NE
BR-NO
BR-SE
PR
RS
SC
UY-MD
UY-RI
UY-TT
BR-UY-II
RI-I
BR-RS-I
RI-II
BR-UY-I
TT-I
Mar 14
Apr 19
May 26
Jul 2
Aug 7

**Figure S2.** Spatiotemporal dissemination of SARS-CoV-2 B.1.1.33 Uruguayan-Brazilian variants. Time-scaled bayesian phylogeographic MCC tree. Branches are colored according to the most probable location state of their descendant nodes as indicated at the legend. Posterior probability/Posterior state probability support values are indicated at key nodes.

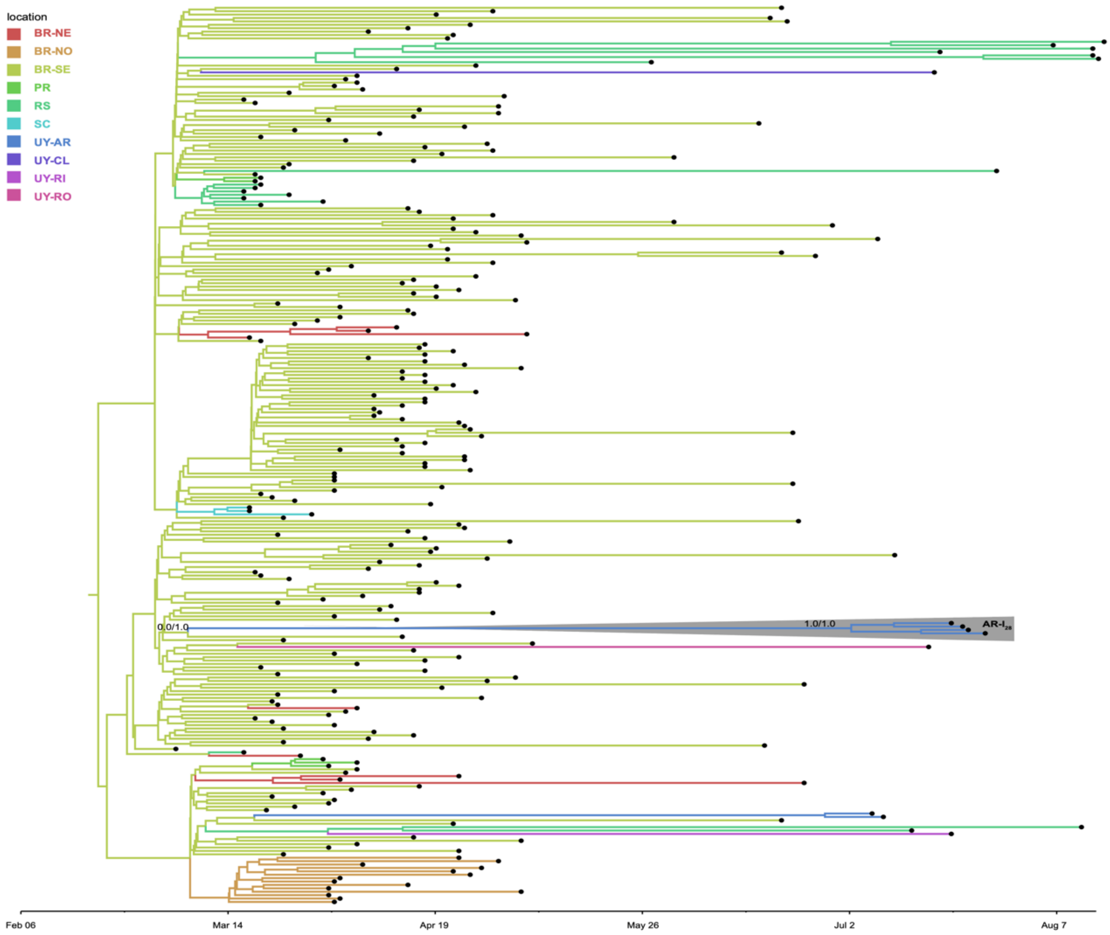

**Figure S3.** Spatiotemporal dissemination of SARS-CoV-2 B.1.1.28 Uruguayan-Brazilian variants. Time-scaled bayesian phylogeographic MCC tree. Branches are colored according to the most probable location state of their descendant nodes as indicated at the legend. Posterior probability/Posterior state probability support values are indicated at key nodes.

[2] dx.doi.org/10.17504/protocols.io.bfy7jpzn, BioRxiv

[3] https://dx.doi.org/10.17504/protocols.io.bdp7i5rn

[4] https://doi.org/10.1038/nprot.2017.066

[5] https://doi.org/10.1038/nprot.2017.066

[6] https://www.biorxiv.org/content/10.1101/2020.10.06.328328v1

[7] https://artic.network/ncov-2019/ncov2019-bioinformatics-sop.html, accessed on December 2020.

[8] https://www.nature.com/articles/nbt.3820

[9] https://github.com/iferres/ncov2019-artic-nf

[10] https://doi.org/10.1038/nmeth.1923

[11] https://doi.org/10.1093/nar/gkf436

[12] doi:10.1093/molbev/msx149

[13] doi: 10.1093/ve/vew007

[14] doi:10.1093/molbev/msi103

[15] doi: 10.1093/ve/veaa061

[16] https://virological.org/t/time-dependence-of-sars-cov-2-substitution-rates/542

[17] doi:10.1093/sysbio/syy032

[18] doi: 10.1002/j.1538-7305.1948.tb01338.x

[19] R Core Team (2020). R: A language and environment for statistical computing. R Foundation for Statistical Computing, Vienna, Austria. URL https://www.R-project.org/
